# Leveraging deep-learning on raw spirograms to improve genetic understanding and risk scoring of COPD despite noisy labels

**DOI:** 10.1101/2022.09.12.22279863

**Authors:** Justin Cosentino, Babak Behsaz, Babak Alipanahi, Zachary R. McCaw, Davin Hill, Tae-Hwi Schwantes-An, Dongbing Lai, Andrew Carroll, Brian D. Hobbs, Michael H. Cho, Cory Y. McLean, Farhad Hormozdiari

## Abstract

Chronic obstructive pulmonary disease (COPD), the third leading cause of death worldwide, is highly heritable. While COPD is clinically defined by applying thresholds to summary measures of lung function, a quantitative liability score has more power to identify new genetic signals. Here we train a deep convolutional neural network on noisy self-reported and ICD-based labels to predict COPD case/control status from high-dimensional raw spirograms and use the model predictions as a liability score. The machine-learning-based (ML-based) liability score accurately discriminates COPD cases and controls (AUROC = 0.82 *±* 0.01) and COPD-related hospitalization (AUROC = 0.89 *±* 0.01) without any domain-specific knowledge. Moreover, the ML-based liability score is associated with overall survival (Hazard ratio = 1.22 *±* 0.01; P ≤ 2 *×* 10^−16^) and exacerbation events (*R*^2^ = 0.10 *±* 0.01; P ≤ 4 *×* 10^−101^). A genome-wide association study on the ML-based liability score replicates existing COPD and lung function loci, but also identifies 67 new loci. Thirty-eight of these have supportive evidence in independent datasets, including a locus near *LTBR*. We demonstrate the biological plausibility of the novel variants through enrichment analyses, phenome-wide association studies, and generalizability of COPD prediction in multiple datasets. These results provide an example of the potential to improve genetic discovery of disease-relevant variants by training deep neural networks to predict noisy labels from high-dimensional raw data.

## 1 Introduction

Chronic obstructive pulmonary disease (COPD) is a lung disorder characterized by impeded airflow and persistent airway inflammation [1]. According to the World Health Organization’s latest assessment, in 2019 COPD was the third leading cause of death world-wide, and the seventh leading cause of disability-adjusted life years (DALYs) [2]. In 2019 alone, 3.2 million deaths and nearly 74 million DALYs were attributed to COPD [2]. Although smoking is a major risk factor, COPD is a complex and heterogeneous disease, with both environmental and genetic components [3, 4]. Among individuals with the same smoking history, not all will go on to develop COPD for reasons that may relate to genetic predisposition [5]. Twin studies and genome-wide analyses have estimated the heritability of COPD at 40–60% [6, 7].

COPD has a consensus definition based on symptoms and spirometry [5]. Spirometry is a quantitative pulmonary function test that measures the volume and rate of air expelled from the lungs. The key summary measures extracted from spirograms for COPD are forced vital capacity (FVC; the total volume of air forcibly expelled starting from maximal inspiration), and forced expiratory volume in 1 second (FEV_1_; the volume expelled in the first second of an FVC maneuver) [8]. Clinically, spirometry summary measures are central to the diagnosis of COPD [5, 9]. According to the Global Initiative for Chronic Obstructive Lung Disease (GOLD) criteria, a post-bronchodilator FEV_1_/FVC ratio less than 0.7 is diagnostic of COPD. Among patients with FEV_1_/FVC *<* 0.7, the severity of airflow limitation in COPD may be graded by comparing observed FEV_1_ with predicted FEV_1_ based on a patient’s age, height, sex, and ethnicity (FEV_1_%predicted).

In recent years, large population biobanks, including the UK Biobank [10], have enabled genome-wide association studies (GWAS) of COPD [11, 12] and lung function [13, 14] that include hundreds of thousands of subjects, based on FEV_1_, FVC, and their ratio. However, GWAS of binary case/control COPD status based on spirometry summary measures may lack power to identify the underlying genetic variants due to the following factors. First, the binary “control” status does not differentiate between normal and pre-COPD patients who do not meet the spirometric cutoffs but might show accelerated lung function decline or be smokers showing significant symptoms and chest imaging abnormalities [15, 16]. This known limitation has led to proposals for new COPD diagnostic criteria not based solely on lung function [17, 18]. Second, the binary “case” status does not capture variation in severity of airflow limitation or frequency of acute exacerbations across patients [5]. Third, COPD is a heterogeneous disease [1, 5], with multiple underlying pathobiologic processes, and different genetic variants likely underlie different processes [19]. This heterogeneity exacerbates the effect of lack of power. Fourth, while analysis of the underlying quantitative trait is likely more powerful than analysis of binary COPD labels, studies of quantitative summary measures of lung function (e.g., FEV_1_ and FVC) may miss specific combinations of measurements or patterns that are more representative of clinically apparent COPD.

In this work, we hypothesized that using raw spirograms to define a COPD liability score would improve power to elucidate the genetic architecture of COPD. We use COPD liability score and COPD risk score interchangeably. Raw spirograms likely contain additional information beyond that captured by common summary metrics and fixed cutoffs, and this information may be relevant to assessing disease risk and severity. To study this hypothesis, we applied deep learning and extended the machine-learning-based (ML-based) phenotyping methodology [20] to predict COPD liability. In ML-based phenotyping, an ML model is used to define a synthetic phenotype whose genetic basis is studied. In previous works [20, 21], it was shown that training ML-based phenotyping models on accurate labels increases association power by providing a continuous metric of disease risk rather than a binary case/control status. However, finding accurate and clinically-graded disease status labels for training ML models can be challenging, expensive, and time-consuming for certain hard-to-collect diseases and disorders. In contrast, partial medical records and self-reported labels are more accessible and can be used to define disease labels despite not being as accurate as clinically defined labels. In this work, we demonstrate that ML-based phenotyping does not necessarily require accurate labels. In particular, ML-based phenotyping models defined on noisy labels, extracted from partial medical records without expert medical review, can provide biologically interesting and clinically predictive phenotypes.

We developed an ML model that utilizes complete spirograms of a single blow, e.g., the entire volume-time curve, trained on medical-record-based binary COPD status labels. In this paper, medical-record-based COPD is defined solely based on self-reporting or the existence of COPD ICD codes in the partial medical records collected by the UK Biobank from primary care and hospital inpatient records. Consequently, medical-record-based COPD is a noisy measure of COPD status. Moreover, in contrast to a case/control label, the proposed model translates a patient’s COPD status into a continuous liability score. Notably, this ML-based liability score is a better predictor of future hospitalization or death primarily caused by COPD than FEV_1_/FVC or FEV_1_%predicted. GWAS for the ML-based COPD risk not only replicates most known COPD associations [12], but identifies 265 additional risk loci; of these, 101 replicate in Sakornsakolpat et al.’s [12] COPD GWAS after Bonferroni correction and 198 were previously identified (i.e., associated) for lung function. For the remaining 67 loci out of 265, we observed that 27 had at least nominal evidence of replication in at least one independent dataset of either COPD or lung function. We further validate and interrogate the GWAS results using multiple post-GWAS analyses, including gene-set enrichment analysis, out of sample polygenic risk prediction, and phenome-wide association studies.

## 2 Results

### 2.1 ML-based COPD phenotyping overview

We developed a deep learning model to predict COPD risk (i.e., an ML-based liability score; Figure **1**) from the spirogram of a single blow (Figure **2**, Supplementary Figures **1** and **2**). The UK Biobank obtained raw volume-time spirograms from almost all participants, and created summary measures including FEV_1_ and FVC from them. We derived flow-time and flow-volume curves from the volume-time curve (Methods).

**Fig. 1:**
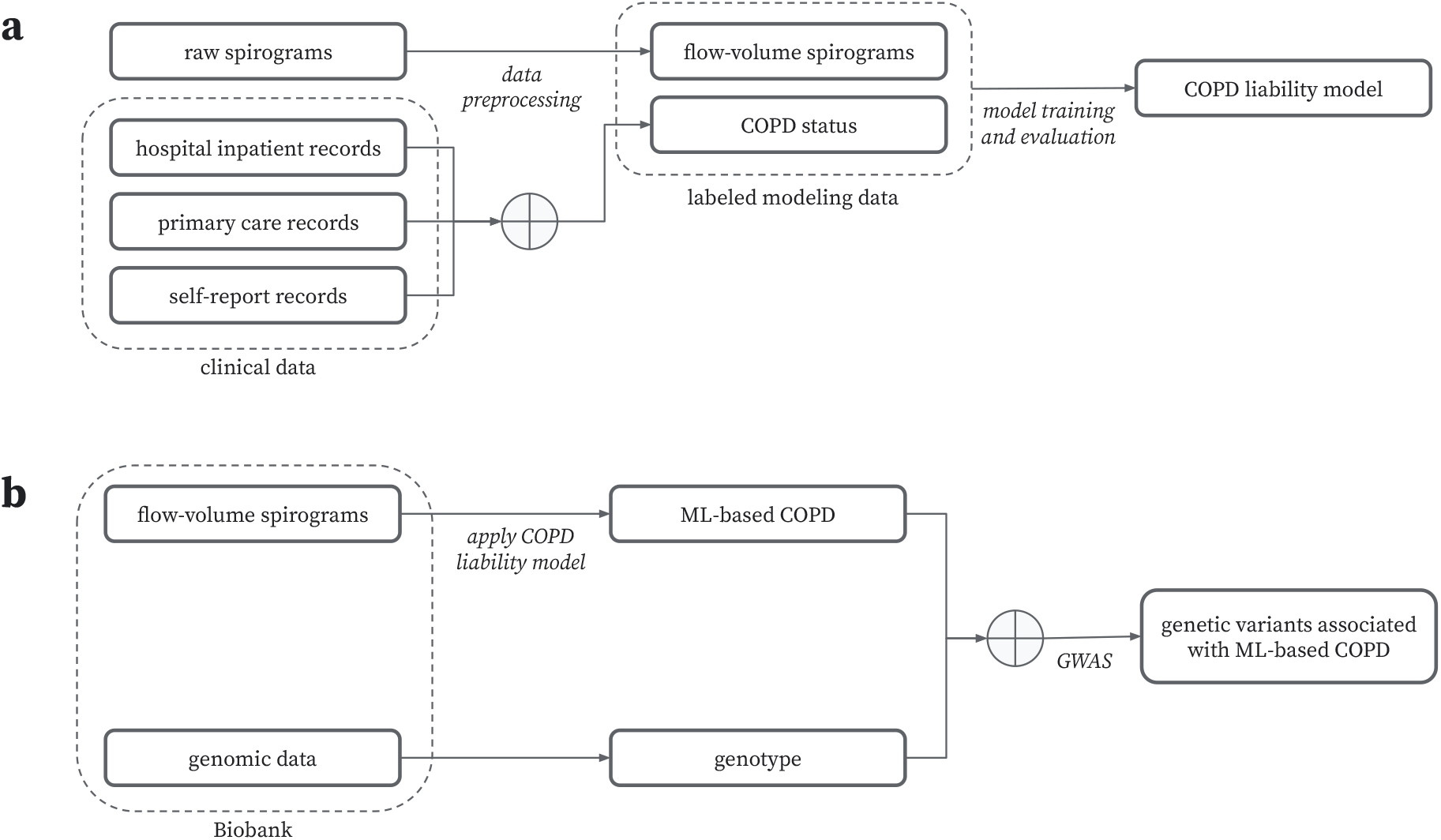
ML-based COPD phenotyping overview. a) During the “model training” procedure, noisy COPD status labels were derived using various medical record sources. A COPD liability model is then trained to predict COPD status from flow-volume spirograms. b) During the “model application” procedure, we applied this COPD liability model to the target cohort’s flow-volume spirograms to generate ML-based COPD liability scores. These liability scores were then paired with genotype data for genomic discovery.

**Fig. 2:**
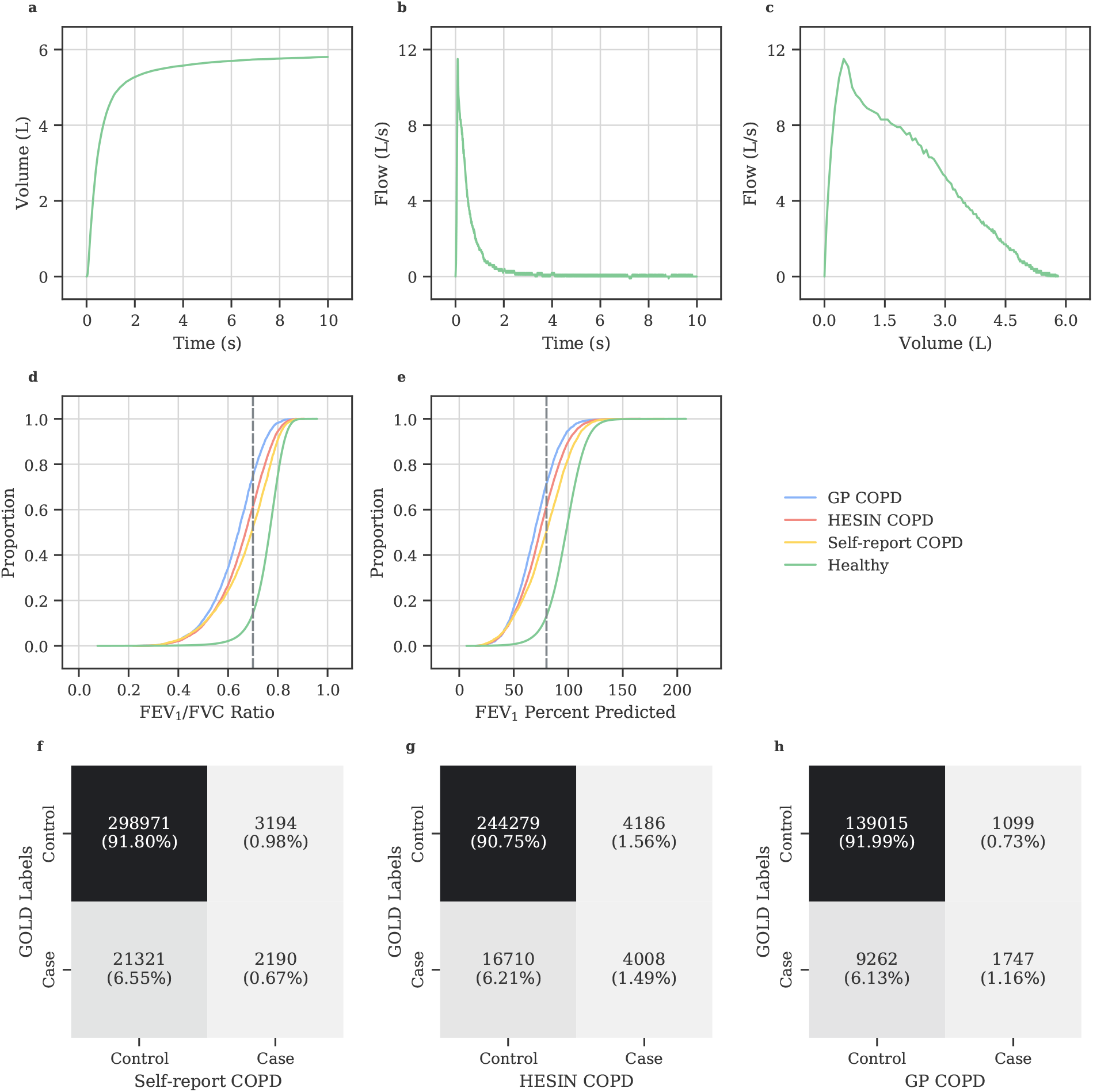
Spirometry and COPD status overview. a) A forced expiratory volume-time spirogram. b) A forced expiratory flow-time spirogram. c) An interpolated forced expiratory flow-volume spirogram. d) A cumulative distribution function showing FEV_1_/FVC ratios of valid spirometry blows in UKB grouped by COPD label source. The dotted line denotes the 0.7 GOLD criteria cutoff for COPD diagnosis. e) A cumulative distribution function showing FEV_1_%predicted of valid spirometry blows in UKB grouped by COPD label source. The dotted line denotes the 80% GOLD criteria cutoff for COPD 2-4 diagnosis. f-h) Confusion matrices for COPD diagnosis between proxy GOLD 2-4 criteria and medical-record-based labels from self-report (f), HESIN (g), and GP data sources (h).

To train our ML model, we used binary COPD labels defined based on partial medical records and self-reported data. We defined “medical-record-based” COPD as self-reported COPD or the existence of any COPD-related ICD code in the partial medical records collected by the UK Biobank through linkages to a range of primary care, referred to as the GP dataset, and hospital inpatient admission records, referred to as the HESIN dataset (see Supplementary Table **1** for the exact definitions). We trained an ensemble of one-dimensional convolutional neural networks (CNNs) [22, 23] based on the ResNet18-D architecture [24, 25] to predict medical-record-based COPD status from first visit flow-volume spirograms. Starting from a volume-time curve consisting of 1,000 points (Figure **2**a), we derived flow-time and flow-volume curves (Figure **2**b and Figure **2** and Methods).

We applied our trained model to generate ML-based liability scores for all unrelated European subjects with acceptable spirograms (*n* = 325, 027, Methods) and then performed GWAS on the predicted liability scores (Figure **1**). Unlike previous ML-based phenotyping works [20, 21, 26] that use high quality labels from experts, our approach uses noisy COPD status from self-reporting and ICD-based partial medical records. These self-reports are expected to be error-prone and, in medical records, undiagnosed or misreported individuals with COPD are common [27]. To estimate the noise in these labels, we compared them against single-blow “proxy-GOLD” labels. We define proxy-GOLD similarly to the GOLD standard for COPD of at least moderate severity: we labeled subjects as likely COPD cases if, for a single blow, their FEV_1_/FVC measurement was *<* 0.7 and their observed FEV_1_ was *<* 80% of their corresponding predicted FEV_1_ value (Methods). Although these labels are not strictly GOLD labels–since a single blow is used and bronchodilation was not applied prior to the spirometry test–they provide a strong measure of COPD status [12, 14]. Similar to previous work [27], we observed that a non-negligible proportion of subjects who met this proxy-GOLD criteria were labeled as a control in medical-record-based COPD (Figure **2**f-h). It is worth mentioning that we could not use proxy-GOLD as labels to our ML model since proxy-GOLD relies on FEV_1_/FVC and FEV_1_, which are directly defined from the spirograms. Thus, utilizing proxy-GOLD in our ML model would result in double dipping of data, creating an undesirable feedback loop (see Discussion).

### 2.2 ML methods improve COPD detection relative to spirometry metrics

In the context of model evaluation for the ML-based liability score, we use three sets of labels (see Supplementary Table **1** for exact definitions): evaluation medical-record-based COPD, defined as above but restricted to the subset of individuals with data available from *all* three data sources (*n* = 125, 786), a future hospitalization indicator, and a mortality indicator. The latter two labels were defined to identify patients with COPD as a primary cause of hospitalization or death, respectively, after their date of spirometry assessment. Note that for evaluation medical-record-based COPD, having data from all three sources increases the likelihood of a correct label, which is preferred for evaluation. We randomly split European-ancestry individuals with acceptable blows into training and validation sets containing 80% and 20% of samples, respectively, to form the “modeling” dataset used to tune model hyperparameters and evaluate the ML-based liability score (section 4.3 and Supplementary Figure **3**).

We trained models based on one-dimensional variants of the multilayer perceptron, LeNet5 [22], ResNet9 [24], and ResNet18 [24] model architectures, and considered multiple representations of the raw volume-time, flow-time, and flow-volume spirograms as inputs, as well as a wide range of hyperparameters specific to each architecture class (Methods and Supplementary Table **2**-**7**). We observed that a ResNet18 model trained using only the flow-volume curve outperforms all other models, including other spirogram-based and spirometry-metric-based ML approaches, across tasks in the validation dataset (Methods and Supplementary Table **3** and Table **4**). Additionally, the flow-volume ResNet18 model outperforms risk scores based on FEV_1_/FVC ratio and FEV_1_%predicted (Figure **3**). Specifically, when compared to FEV_1_%predicted, which often outperforms FEV1/FVC ratio (Supplementary Table **3**), the flow-volume ResNet18 shows improved AUROC and AUPRC predictive performance for medical-record-based COPD status (AUROC = 0.82 *±* 0.01 vs. 0.78 *±* 0.01, AUPRC = 0.33 *±* 0.03 vs. 0.21 *±* 0.02; Figure **3**a and d), future COPD-related hospitalization (AUROC = 0.89 *±* 0.02 vs. 0.87 *±* 0.02, AUPRC = 0.18 *±* 0.03 vs. 0.09 *±* 0.02; Figure **3**b and e), and COPD-related death (AUROC = 0.95 *±* 0.03 vs. 0.92 *±* 0.05, AUPRC = 0.06 *±* 0.03 vs. 0.03 *±* 0.02; Figure **3**c and f). Paired bootstrapping over *n* = 100 trials showed that all of these differences are significant (Supplementary Table **4**).

**Fig. 3:**
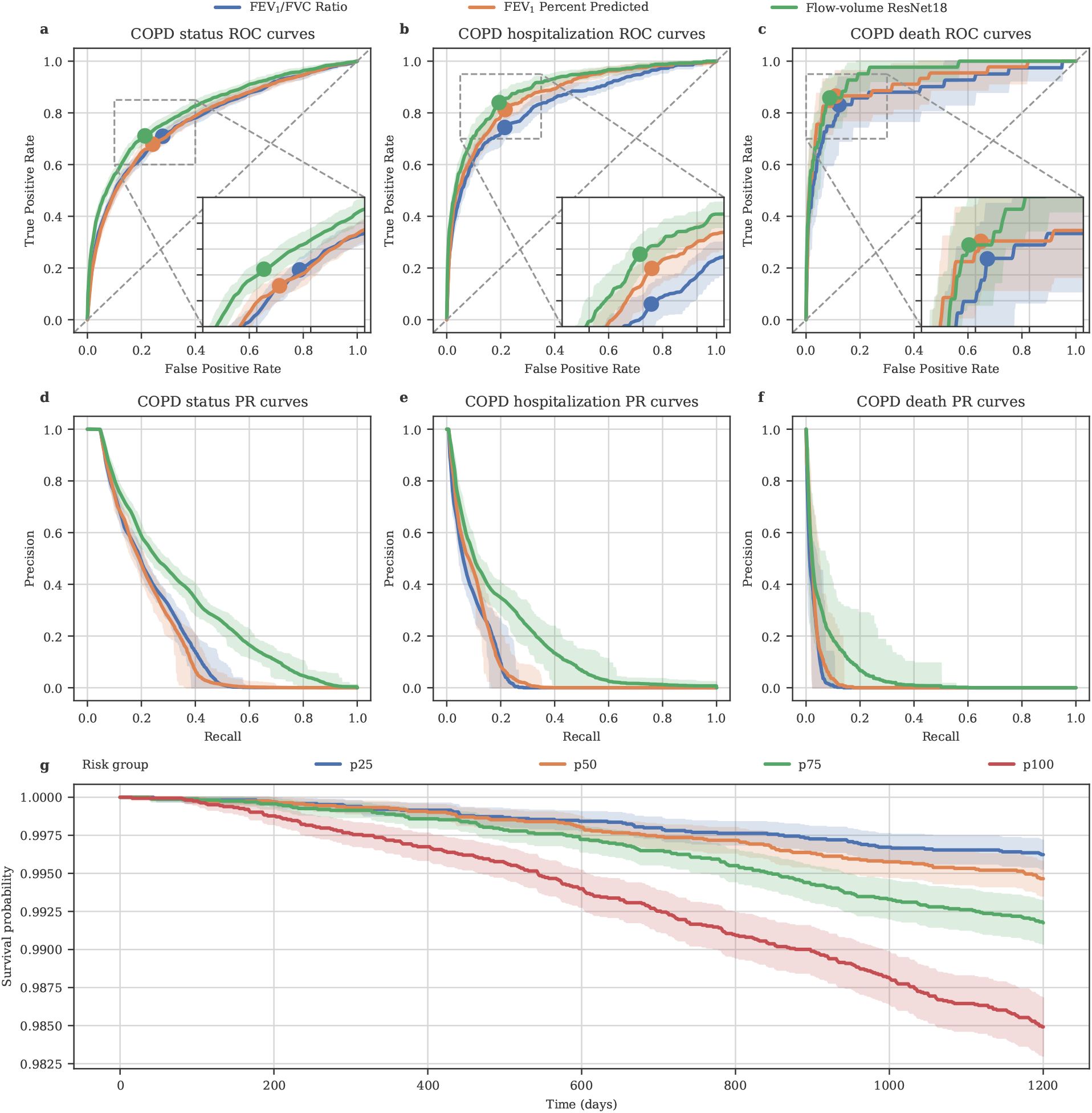
ML methods improve COPD detection relative to spirometry metrics in the UKB modeling validation set. a-c) A comparison of ML-based COPD risk, FEV_1_/FVC-ratio-based risk, and FEV_1_%predicted-based risk receiver operating characteristic (ROC) curves across the evaluation medical-record-based COPD (left), future COPD-related hospitalization (center), and COPD-related death (right) tasks. Error bars denote bootstrapped 95% confidence intervals (*n* = 100 bootstrapping samples). d-f) A comparison of flow-volume ResNet18 COPD predictions, FEV_1_/FVC-ratio-based risk, and FEV_1_%predicted-based risk precision-recall (PR) curves across the evaluation medical-record-based COPD (left), future COPD-related hospitalization (center), and COPD-related death (right) tasks. Error bars denote bootstrapped 95% confidence intervals (*n* = 100 bootstrapping samples). g) Kaplan-Meier curves estimating the survival function of individuals grouped into quartiles by ML-based COPD risk.

### 2.3 ML-based COPD risk is associated with survival time and exacerbation

We performed survival analysis using the aforementioned COPD-related mortality labels for all individuals in the *modeling* dataset’s validation split (*n* = 65, 281, Supplementary Figure **3** Section 4.3), fitting a Cox proportional hazards regression model to UKB death registry data while controlling for age and sex as covariates (Methods). The hazard ratio (HR) for *all* death (i.e., overall survival) was 1.22 (SE = 0.01; P ≤ 2 *×* 10^−16^) per one standard deviation (1-SD) increase in ML-based liability score. Kaplan-Meier curves for overall survival (OS) stratified by ML-based liability score indicate that OS declines more rapidly for patients at higher COPD liability (Figure **3**g). Furthermore, using COPD-related hospitalization (HESIN) episodes as a proxy for COPD exacerbation, we observed that ML-based liability score is significantly better correlated with an individual’s number of exacerbatory events (*R*^2^ = 0.1031 *±* 0.0110; P ≤ 4 *×* 10^−101^) when compared to FEV_1_/FVC ratio (*R*^2^ = 0.0370 *±* 0.0037) and FEV_1_%predicted (*R*^2^ = 0.0285 *±* 0.0030) spirometry metrics (Supplementary Table **5** and Table **6**).

### 2.4 ML-based COPD captures 265 novel association loci

We generated ML-based liability scores for all European ancestry individuals with acceptable spirograms in UK Biobank (*n* = 325, 027). To maximize the accuracy of this ML-based phenotyping procedure and avoid data leakage, the ML-based liability scores used in GWAS were obtained by 2-fold cross validation. A separate model was trained on each fold and then applied to the other fold to generate liability scores (Supplementary Figure **4**; see Methods). The folds were constructed to keep genetically related individuals together, preventing the same individual or a close relative from being used for both training and prediction. We investigated the effect of sample size and 2-fold cross validation on performance. An ablation study examining the impact of training dataset size on ResNet18 model performance (Supplementary Figure **5**) and a comparison of cross-fold model predictions (Supplementary Table **7**) show that performance is consistent under the 2-fold cross-validation approach (Methods).

We performed GWAS on the ML-based liability scores using BOLT-LMM [28] adjusting for age, sex, age *×* sex (age and sex interaction), genotyping array, standing height, standing height *×* standing height (standing height squared), body mass index (BMI), smoking status, and the top 15 genetic principal components (PCs) as covariates (Methods). To improve the statistical power of the GWAS, we applied a direct inverse normal transform (D-INT) [29] to the ML-based liability scores (Figure **4**a). Although the genomic inflation *λ*_*GC*_ was 1.49 (Supplementary Figure **6**), the stratified linkage disequilibrium score regression-based (S-LDSC) [30] intercept was only 1.07 (s.e.m = 0.02), indicating that the inflation of *λ*_*GC*_ is attributable to high polygenicity rather than confounding or population structure. The SNP-heritability estimated from S-LDSC for ML-based COPD was 0.20 (s.e.m = 0.01). The ML-based COPD GWAS identified 796 independent genome-wide significant (GWS) hits (*R*^2^ ≤ 0.1 and P ≤ 5 *×* 10^−8^; Supplementary Table **8** and Supplementary Table **9**) at 356 independent GWS loci after merging hits within 250kb together (Supplementary Table **10**). Of these, 433 hits (Supplementary Table **11** and Table **12**) within 265 loci (Supplementary Table **13** and Table **14**) have not previously been associated with COPD.

**Fig. 4:**
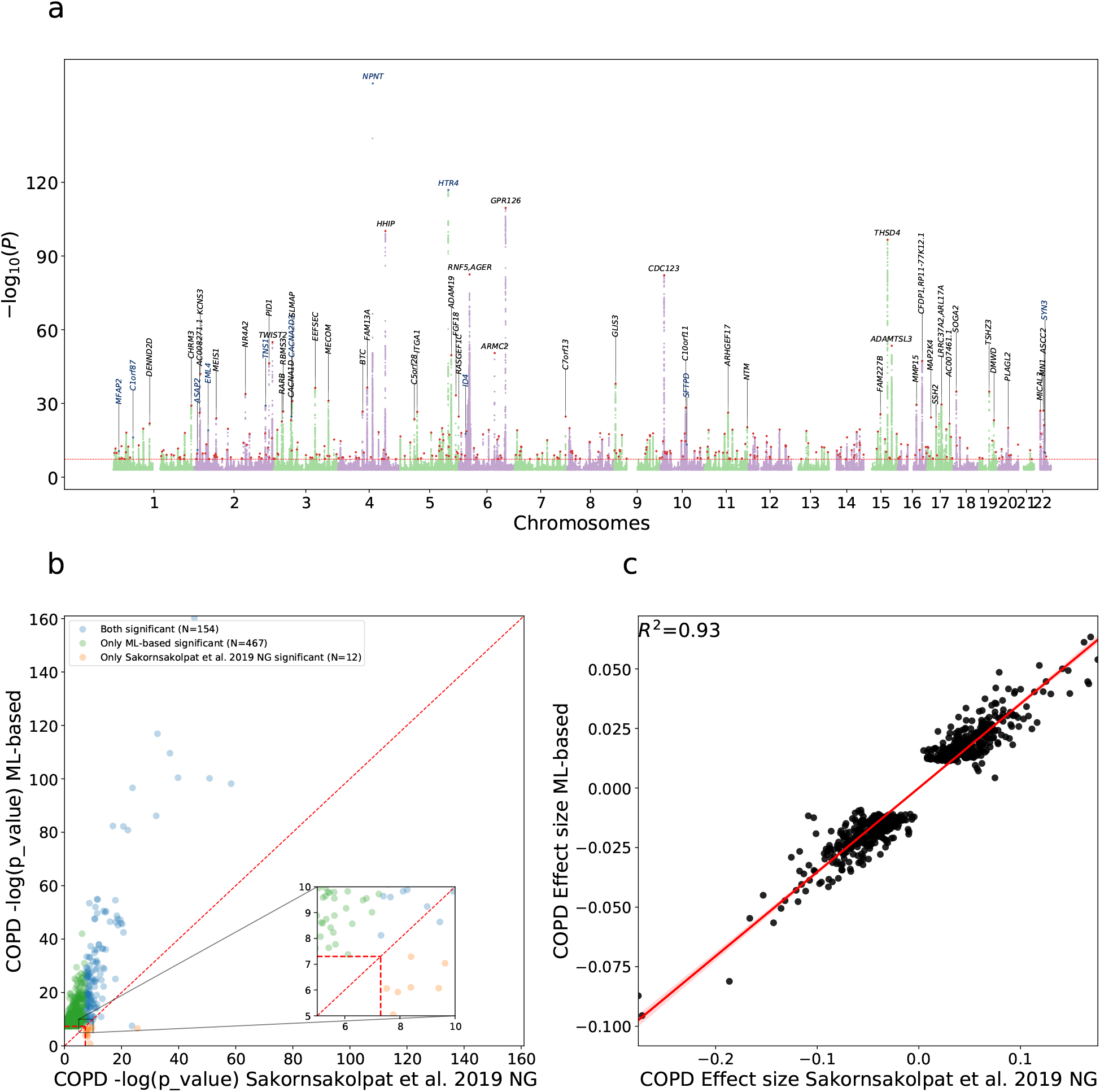
ML-based COPD captures 266 novel association loci. a) Manhattan plot depicting ML-based COPD-associated GWAS p-values for all 22 autosomal chromosomes. Black gene names indicate the closest gene for each locus with log_10_ *p >* 20 and red dots denote all other GWS loci. Blue gene names and dots indicate loci also identified in the Sakornsakolpat et al. [12] study. Supplementary Table **10** contains a complete list of all GWS loci. b) Comparison of ML-based significance level GWS hits with existing COPD GWAS of Sakornsakolpat et al. [12]. The X-axis is the− log p-value of Baseline (Sakornsakolpat et al. [12]). The Y-axis is the − log p-value of the ML-based COPD. Both p-values are computed using two-sided tests. The vertical and horizontal red lines indicate the genome-wide significance level. The diagonal red line indicates *y* = *x*. The orange dots indicate variants that are significant for Baseline (Sakornsakolpat et al. [12]) but not significant for our ML-based COPD and green dots indicate variants that are significant for our ML-based COPD but not significant for Baseline. c) Effect size correlation of ML-based COPD and Baseline (Sakornsakolpat et al. [12]) COPD GWAS. The X-axis is the effect size of Baseline COPD for all GWS hits and Y-axis is the effect size of our ML-based COPD.

Previous works [12, 14] showed that many COPD hits are shared with FEV_1_/FVC, FEV_1_, FVC, and peak expiratory flow (PEF) hits. To ensure our ML-based COPD GWAS is not solely driven by FEV_1_/FVC, we performed a secondary ML-based COPD GWAS conditioned on FEV_1_/FVC (Supplementary Figure **7** illustrates the Manhattan plot and Supplementary Figure **8** illustrates the Q-Q plot). The SNP-heritability estimated from S-LDSC for conditional GWAS was 0.11 (s.e.m=0.01). The conditional GWAS identified 175 independent GWS hits at 117 independent GWS loci after merging hits within 250kb (Supplementary Table **15** and Table **16**). Although the conditional analysis ensures that our ML-based COPD GWAS is not solely driven by FEV_1_/FVC, it does not rule out cases where FEV_1_/FVC has a non-linear effect on ML-based COPD. We utilized DeepNull [31] to account for possible non-linear relationships between age, sex, and FEV_1_/FVC. The ML-based COPD GWAS using DeepNull (Supplementary Figure **10** illustrates the Manhattan plot and Supplementary Figure **11** illustrates the Q-Q plot) identified 181 independent GWS at 129 independent GWS loci after merging hits within 250kb (Supplementary Tables **19** and **20**). Thus, our ML-based COPD prediction captures a disease signal beyond FEV_1_/FVC (Supplementary Table **21** and Table **22**). Furthermore, we performed ML-based COPD GWAS conditioned on FEV_1_/FVC, FEV_1_, FVC, and PEF, observing a SNP-heritability of 0.04 (s.e.m = 0.00) and 41 independent GWS hits at 31 independent GWS loci (Supplementary Figure **9**, Supplementary Table **17** and Table **18**).

Finally, to ensure that this result is not influenced by some bias introduced by our ML-based phenotyping procedure, we trained the ResNet18 model using permuted medical-record-based COPD labels and observed that the model predicts almost the same value for all individuals (0.0384*±*0.0000), which matches the disease prevalence in the training data. In other words, the model cannot detect any patterns from inputs to the permuted labels and falls back on the best guess of prevalence for the probability of having COPD. Furthermore, we ran a GWAS on a permuted version of the original ML-based COPD phenotype and observed that this permuted GWAS has SNP-heritability of zero (0.00 *±* 0.01) and that no GWS variants were detected.

### 2.5 ML-based COPD improves GWAS statistical power

We compared our ML-based GWAS with the results of the largest available meta-analysis, that from Sakornsakolpat et al. [12]. 220 GWS hits were only significant in the ML-based GWAS while 9 were only significant in the Sakornsakolpat et al. meta-analysis [12]. In addition, as shown in Figure **4**b, most of the GWS hits are shifted toward the ML-based axis indicating that, for common hits, ML-based COPD p-values are smaller than the corresponding Sakornsakolpat et al. values. This suggests that our ML-based GWAS has higher statistical power. The genetic correlation of ML-based COPD prediction and Sakornsakolpat et al. [12] using S-LDSC was *r*_*g*_ = 0.90 (s.e.m = 0.07, Supplementary Table **23**) and the effect size correlation of GWS hits was *R*^2^ = 0.93 (Figure **4**c). Thus, ML-based COPD GWAS appears to improve statistical power by reducing the standard error of effect size estimates compared to the previous work of Sakornsakolpat et al. [12].

There are two potential explanations for improved statistical power: first, utilizing liability scale (i.e., continuous risk) of COPD disease instead of case/control (i.e., binary) and second, the ML-based COPD identifies clinically defined disease slightly better than proxy-GOLD for genetic discovery (perhaps, on the edge cases of fixed cutoffs). All results before this section indicate the former. In this section, we will show that our ML-based COPD risk serves as a better disease liability score than proxy-GOLD for genetic discovery. First, we binarized the ML-based COPD risk into case/control labels with 50% prevalence (Methods) and compared a GWAS on this phenotype (hereafter “binarized ML-based COPD”) with GWAS performed on medical-record-based COPD labels. We observed that binarized ML-based COPD has a higher significance level for all hits (351/354 are only significant in binarized ML-based COPD and 1/354 are significant in both GWAS) and the absolute magnitude of binarized ML-based COPD is larger than the raw label equivalents for all hits (Supplementary Figure **12**). We compared our binarized ML-based COPD with Sakornsakolpat et al. [12] where we observed that our binarized ML-based COPD variants are more significant (Supplementary Figure **13**a) than Sakornsakolpat et al. [12] while having the same effect size estimates (*R*^2^ = 0.91; Supplementary Figure **13**b). Furthermore, when binarizing ML-based COPD so that disease prevalence matches Sakornsakolpat et al. [12] (prevalence = 13.86%), the prevalence-matched binarized ML-based COPD has better power. Finally, when comparing a binarized ML-based COPD where we match the prevalence to proxy-GOLD, we observed that binarized ML-based COPD outperforms proxy-GOLD on all metrics including replicating previously known COPD hits (Supplementary Figure **14** and Figure **15**). Thus, even when ML-based COPD is considered as a binary trait, we show increased power (Supplementary Table **8**).

A GWAS on the ML-based liability score identifies 265 novel COPD risk loci in addition to 91 previously known COPD loci with respect to to Sakornsakolpat et al. [12] and GWAS catalog entries (as of 2022-07-09) for COPD, emphysema, chronic bronchitis. Out of 265 novel loci, 221 independently replicate as associated with COPD or COPD-related lung function as follows. We observed that 101 out of 265 replicate in a previous COPD GWAS [12] after Bonferroni correction. Also, 198 out of 265 are previously known FEV_1_ or FEV_1_/FVC loci with respect to [14] and GWAS catalog entries (Supplementary Table **24** and Supplementary Figure **16**). From the remaining 67 loci out of 265, which are not previously known loci for COPD or COPD-related lung function, 23 replicate in a previous COPD GWAS [12] after Bonferroni correction that includes UK Biobank samples. Furthermore, we analyzed three additional studies that do not include UK Biobank samples to further quantify the replication status of these 67 loci. These three datasets are GBMI (Global Biobank Meta-analysis Initiative) [32], SpiroMeta [33], and ICGC (International COPD Genetics Consortium) [11]. We defined two replication strategies: First, we defined *supportive* replication as consistent effect size direction between our ML-based COPD and the three comparators. The ICGC and GBMI GWAS are based on a COPD phenotype; thus, we expect their effect size signs to match our ML-based COPD. SpiroMeta phenotypes, on the other hand, capture lung function, so we expect their effect size signs to be the opposite of our ML-based COPD signs. Second, we defined *strict* replication as consistent effect size direction in any study with Bonferroni-corrected P < 0.1 (one-sided) for that study. We observed that 38/67 loci have *supportive* replication where the chance of this happening randomly is extremely small (P ≤ 2 *×* 10^−16^). In addition, we observed that 6/67 have *strict* replication and, when relaxing the strict replication p-value from Bonferroni-corrected to nominal P *<* 0.1, 27/67 loci replicate (Supplementary Table **24** and Supplementary Figure **16**).

### 2.6 ML-based COPD enriched in lung tissue

Utilizing S-LDSC to perform tissue and cell-type specific analysis, we observed that fetal lung and smooth-muscle are the relevant tissues for ML-based COPD (Supplementary Tables **25** and **26**). These tissues and cells are similar to previous work [12], further indicating that our ML-based COPD is a valid COPD phenotype and the improvement observed in number of additional hits/loci are not due to capturing other non-COPD phenotypes with high heritability (e.g., height). Furthermore, we observed that colon smooth muscle (H3K4me1; P = 5 *×* 10^−10^) and fetal lung (H3K4me1; P = 2 *×* 10^−9^) are the relevant tissues for ML-based COPD GWAS conditional on FEV_1_/FVC (Supplementary Table **27**). Similar to S-LDSC analysis, GARFIELD [34] indicated that the fetal lung has the largest enrichment (Supplementary Figure **17**) and the conditional GWAS of ML-based COPD on FEV_1_/FVC was enriched in fetal lung and embryonic lung (Supplementary Figure **18**). Lastly, to understand the effect of cis-regulatory interactions, we applied GREAT [35] to ML-based COPD GWS loci. The ML-based loci were significantly enriched for 82 ontology terms, primarily development and morphogenesis-related. Of particular note, GREAT results were enriched for respiratory and cardiovascular system development and morphology terms (Supplementary Table **28**).

### 2.7 ML-based COPD hits detect high risk COPD cases

To evaluate the quality of hits, we examined their collective predictive power for detecting high risk COPD cases by combining them into a simple polygenic risk score (PRS). We observed that the ML-based COPD hits detect high risk COPD cases in both UKB and COPDGene [36]. We compared this PRS with the PRSs defined based on the hits of the medical-record-based COPD and Sakornsakolpat et al. [12] GWASs. These simple GWAS PRSs are defined on the index variants of hits by multiplying the number of effect alleles by the effect size of a variant (Methods). The main purpose of this experiment is to evaluate the quality of hits and index variants themselves and we did not search for the best PRS possible. We evaluated these hit groups and their equivalent simple PRSs on a holdout set in UKB and cross-dataset on the COPDGene.

The ML-based PRS detects high risk COPD cases in a holdout set in UKB. The holdout set is from the European individuals who are not used in the ML modeling and in the GWASs (*n* = 110, 739). We evaluated the AUROC of these PRSs on three groups of binary outcomes (Supplementary Table **1**): 1) evaluation medical-record-based COPD, 2) being hospitalized with COPD as the primary cause, and 3) death because of COPD as the primary cause. The results are presented in Table **1**. The ML-based PRS detects high risk COPD cases. Also, it is significantly better than the PRS of the medical-record-based COPD (i.e., the labels on which the model was trained) and Sakornsakolpat et al. [12] when evaluated on the medical-record-based COPD and hospitalization and better (not statistically significant) on the COPD death (where we have small number of deaths). Finally, we evaluated the PRS of a conditional ML-based COPD GWAS where FEV_1_/FVC was one of the covariates and observed even this PRS detects high risk cases with an AUROC of 0.525 (95% CI, 0.515 – 0.534). We observed the same trends when evaluated on AUPRC, top decile prevalence, and Pearson correlation (Supplementary Table **29**).

**Table 1:** ML-based COPD PRS detects high risk COPD cases. MRB stands for medical-record-based. The PRSs are defined based on the GWAS effect sizes of ML-based COPD, medical-record-based COPD, and Sakornsakolpat et al. [12]. The reported metric is AUROC where the numbers in the parenthesis show the 95% confidence interval. The PRSs are compared on UKB holdout set and COPDGene. The UKB holdout set is not used in the GWASs or ML modeling. In COPDGene, the effected individuals are defined as the individuals with final GOLD stage 2, 3, and 4 post-QA. Bold values indicate statistical significance of ResNet18 PRS compared to others.

| Dataset | Ground Truth | Prevalence | ResNet18 PRS | MRB PRS | Sakornsakolpat et al. PRS |
| --- | --- | --- | --- | --- | --- |
| UKB | Evaluation MRB COPD | 0.075 (3351/44780) | **0.550 (0.541–0.560)** | 0.517 (0.509–0.528) | 0.538 (0.529–0.548) |
| UKB | Hospitalization | 0.018 (1731/97977) | **0.564 (0.549–0.577)** | 0.514 (0.504–0.526) | 0.551 (0.537–0.565) |
| UKB | Death | 0.002 (237/110739) | 0.598 (0.557–0.632) | 0.503 (0.473–0.537) | 0.575 (0.533–0.606) |
| COPDGene | COPD status | 0.528 (3471/6576) | 0.615 (0.598–0.631) | 0.525 (0.511–0.538) | 0.616 (0.599–0.630) |

The ML-based PRS also detects high risk COPD cases in COPDGene. We defined a binary outcome for COPD status as the European individuals having GOLD stage 2, 3, or 4 (i.e., GOLD stage 2 and greater). Note that the COPD definition here is GOLD-based and different from the medical record-based definitions in UKB, which reinforces the robustness of the results. We evaluated the AUROC of the three PRSs (Table **1**). The ML-based and Sakornsakolpat et al [12] PRSs have equivalent performance and they were both better than medical-record-based COPD PRS. We observed the same trends when evaluated on AUPRC, top decile prevalence, and Pearson correlation (Supplementary Table **30**). We also measured the Pearson correlation of these PRSs with two quantitative Computed Tomography-based phenotypes. The correlations with “emphPc” (percentage of low attenuation areas <-950 Hounsfield units) are 0.110 (95% CI, 0.085 – 0.135) and 0.140 (95% CI, 0.113 – 0.168) for ML-based and Sakornsakolpat PRSs, respectively. The correlations with “Pi10” (the square root of the wall area of a hypothetical airway with internal perimeter of 10mm) are 0.047 (95% CI, 0.024 – 0.068) and 0.021 (95% CI, -0.005 – 0.042) for ML-based and Sakornsakolpat PRSs, respectively. The ML-based PRS has a statistically better correlation with “Pi10”, while Sakornsakolpat PRS has a better correlation with “emphPct” (Supplementary Table **30**).

### 2.8 PheWAS analysis of significant ML-based COPD hits

Phenome-wide association studies (PheWAS) are used to examine pleiotropic effects, which are particularly relevant when considering pharmacological interventions on implicated genes or pathways. We performed PheWAS for the 796 independent ML-based GWAS hits using 4,083 phenotypes in UKB and 2,803 phenotypes in FinnGen. We used a false discovery rate (FDR) of 5% to detect phenotype and variant pairs that are significant in our PheWAS (Supplementary Table **31**). Not surprisingly, most of the significant associations detected by PheWAS are related to different lung function measures, such as FVC, FEV_1_, FEV_1_/FVC, and PEF (Supplementary Table **32**). Similar to Sakornsakolpat et al. [12], our PheWAS analysis identified association with body composition: Weight (131 hits), BMI (96 hits), and fat-free mass (89 hits). In addition, PheWAS detected multiple significant associations with blood counts: white blood cell count (85 hits), red blood cell counts (85 hits), haemoglobin concentration (85 hits), and platelet count (83 hits).

## 3 Discussion

Although thought to be substantial, the genetic component of COPD remains to be fully elucidated, even in the era of Biobank-scale datasets. Power to identify the underlying genetic variants is likely limited by misclassification, use of fixed thresholds for defining COPD status, and failure to differentiate cases according to disease severity. Thus, in this work, we developed an ML model that leverages a patient’s entire spirogram, a time series of the volume of air expelled from their lungs across time, to provide a COPD risk score that has increased power to detect genetic associations and stronger associations with outcomes than standard spirometry measures. Our previous work [20] relied upon high quality disease labels, provided by ophthalmologists, to train an ML model to accurately discern glaucoma risk based on retinal fundus imagery. A key contribution of the present work is the demonstration that an ML model trained on imperfect medical-record-based labels remains effective for assessing disease risk. Moreover, the risk scores generated by this model served as a useful proxy phenotype for genetic discovery. Importantly, the medical-record-based labels used to train this model were derived from self-reporting and hospital billing codes, and did not require domain knowledge or expert curation, which is scarce, expensive, and time-consuming. Our ML-based COPD risk score accurately discriminates COPD cases and controls, and was significantly correlated with COPD-related hospitalization. Interestingly, our ML-based COPD risk score is associated with overall survival and exacerbation events. In the context of genetic discovery, our GWAS of ML-based COPD risk detected 265 novel GWS loci while replicating 221 loci. Lastly, a simple polygenic risk score obtained from ML-based COPD hits is highly informative to distinguish case/control status in UK Biobank and COPDGene (URLs). These results indicate that our proposed ML-model is clinically informative and a useful proxy phenotype for studying the genetic basis of COPD.

Our ML-based COPD GWAS finds additional signals at genome-wide significance. One set of findings likely relates to overlap with asthma. *IL*33 has been previously strongly associated with asthma, and has already shown promise for COPD in clinical trials [37]. Though an association with COPD has been previously reported, this association defined cases using diagnosis codes and without spirometry. In addition, rs752993 (nearest gene, *CHRNA*2) and rs6889076 (*CCNO*) are both associated with eosinophil counts [38]. Interestingly, mutations in *CCNO* are associated with primary ciliary dyskinesia [39, 40] which can manifest as obstructive lung disease, and ciliary dysfunction is implicated in COPD [41, 42]. A second set of findings likely relates to the genetics of smoking, rs13109980 (*MAML*3) and rs4953148 (*SIX*3) have been associated with lifetime smoking [43]. A third set of associations near *BCL*9 (rs17160467) as well as *FZD*3 (rs117746305) and *SFRP* 1 (rs10092045) [44] further solidify the role of Wnt/β-catenin in COPD pathogenesis. A fourth set of loci relate to immune dysfunction, which is strongly hypothesized to play a role in COPD, but to date has limited support from genetic associations. For example, rs5831575 is proximal to *BCL*11*A* - a transcription factor (TF) important for B cells, found to be differentially expressed in *Hhip* knockout mice [45]. Another example is rs10849448 near *LTBR. LTBR* signaling leads to the development of tertiary lymphoid structures in COPD, and blocking *LT* β*R* in an animal model induced regeneration by preventing epithelial cell death and activating WNT/β-catenin in alveolar epithelial progenitors [46]. Finally, we note that recently, a larger GWAS of lung function has been published; some of our novel findings are confirmed by examination in a larger sample size. For example, rs72703234 (near *DMRT* 2) did not meet replication criteria in Shrine et al. [14], but was reported in 2022 [47].

Although not provided by experts, our supervised learning approach still does require a set of labels. We highlight several reasons for electing to use a medical-record-based definition of COPD over proxy GOLD status. First, we wanted our noisy labels to be based on a distinct data modality from the input to the risk prediction model. As GOLD status labels are defined in terms of the FEV_1_ and FVC, which are in turn computed from the spirogram, using GOLD-based labels would create an undesirable feedback loop. Theoretically, given enough input data, the ML model would learn to recapitulate the GOLD criteria. However, this behavior is not useful, as the GOLD criteria are already concrete and easy to implement. Moreover, for a model that has learned to replicate the GOLD criteria, the risk prediction distribution would concentrate near 0 and 1, resulting in poor resolution for differentiating patients. Second, we hypothesized that the full spirogram contains information beyond what is captured by the common summary metrics such as FEV_1_ and FVC. By using the medical-record-based COPD labels, we enable the model to learn any features of the spirogram that might be relevant to COPD diagnosis, rather than encouraging the model to relearn the common summary metrics used by the GOLD criteria. Finally, medical-record-based disease labels may be useful in settings where a clearly defined and broadly accepted disease definition is unavailable. Our ability to train a performant model on labels derived from noisy billing codes suggests that the absence of expert labels does not preclude development of a viable risk prediction model.

We hypothesized that using raw spirograms to define COPD liability would improve power for genetic discovery in COPD and showed results that support this hypothesis. The ResNet18 model that receives full spirograms as input outperforms complex models of spirometry metrics. This suggests that these curves might be underutilized and that there is extra information in the full curves, relevant to COPD, not captured by the common summary metrics (Supplementary Table **4**). Also, while most of the GWAS power increase stems from moving from a binary GOLD-based phenotype to a quantitative liability phenotype, we conjecture that some part of the power increase might be the result of looking at the full curve, which uses information beyond the fixed cutoffs on summary spirometry metrics. This might be inline with recent efforts to explore proposals for new COPD diagnostic criteria [17, 18]. We do not have the data to fully validate this conjecture, but the experiment in which we binarized the liability score to have the same prevalence as single blow proxy-GOLD provides weak support for this idea. In that experiment, with the exact same sample size and number of cases, the binarized ML-based liability replicated more known COPD hits and outperformed proxy-GOLD (Supplementary Figures **14** and **15**, and Supplementary Table **8**).

Our work has several limitations. First, our analyses only include individuals of European ancestry. While the ML model is robust for non-Europeans (Supplementary Table **33**), the small sample size lacks power for a GWAS. Second, the main purpose of our PRS experiment is to evaluate the quality of our hits and index variants themselves and we did not optimize for PRS performance. Thus, creating the best COPD PRS which is transferable to different populations is an active future research direction. Third, because the spirograms present in UK Biobank were obtained without the use of bronchodilation, we cannot strictly adhere to the GOLD COPD criteria. Instead, similar to previous works [12, 14], our proxy-GOLD labels are based on pre-bronchodilation spirometry measurements. Fourth, while some individuals in the UK Biobank had up to three acceptable blows, our risk scoring only makes use of the first acceptable blow. Incorporating information from all acceptable blows on a patient may improve our risk scoring. Fifth, though we performed multiple conditional analyses (e.g., included smoking as a covariate and utilized DeepNull to model non-linearity), some detected loci, such as that near *CHRNA*2, may be due to an association with smoking. Arguably, such loci remain relevant as smoking is an established cause of COPD, and, as demonstrated at the chromosome 15 locus, smoking-associated loci may have complex effects on phenotypes ([48, 49]). Lastly, our risk prediction model is agnostic to COPD-related phenotypes such as BMI, height, and smoking. Thus, it is possible that our model is learning to indirectly infer an individual’s BMI, height, or smoking status as part of the risk prediction process. Even more interesting, we observed that training the ML-based model with these covariates as additional inputs resulted in improved ML task performance but decreased genetic signal in downstream analyses. This suggests that when the covariates are part of the input, the model focuses on non-COPD genetic components more and, in our main model, we are not overfitting to such implicit signals.

Notwithstanding the above limitations, we have demonstrated that a risk prediction model trained on noisy medical-record-based labels can provide clinically predictive and genetically informative risk scores without requiring expert domain knowledge. Due to the widespread and increasing availability of data from electronic health records, this finding significantly expands the set of diseases for which ML-based risk prediction may be possible. Finally, we anticipate that our strategy of leveraging high dimensional data (e.g., an entire spirogram) to generate a continuous risk score will outperform studying binary labels for a wide range of diseases, improving GWAS power and increasing our understanding of biological mechanisms.

## 4 Methods

### 4.1 Spirogram preparation

Raw volumetric flow curves were sourced from UK Biobank field 3066, which contains exhalation volume in milliliters sampled at 10 millisecond intervals. We converted these measures to liters and then computed the corresponding flow curve by approximating the first derivative with respect to time by taking a finite difference. Volume-time and flow-time curves were normalized to length 1,000 by either truncating long curves or by right-padding short curves using the curve’s final value or zero, respectively. Only 13,605 of the 325,027 valid blows in the modeling dataset (Section 4.3) exceeded 1,000 points. The resulting volume-time and flow-time curves are then combined to generate a one-dimensional flow-volume curve (Figure **2**a-c). To ensure that flow points are sampled at consistent intervals across blows, we converted volume-time curves to monotonic representations by accumulating the maximum volume value over time. We then interpolated 1,000 evenly spaced points between 0 and 6.58, the maximum volume value across the modeling dataset blows, from the given flow-monotonic-volume curves.

To quality control the blows, we drop any blow if one of FEV_1_, FVC, and PEF values is in the extreme tail of all observed values (top or bottom 0.5%). The assumption is that these blows are likely to be noisy. We also remove blows that fail the acceptability (i.e., *valid*) provided by UK Biobank. We deem a blow valid if the value recorded in Field 3061 is 0 (i.e., no problems) or 32 (0×20 - “USER_ACCEPTED” i.e., accepted by investigator). When there is more than one acceptable blow, we choose the first one (in the order provided by UK Biobank).

### 4.2 Phenotype definitions

Data were synthesized across several UK Biobank fields to manually define medical-record-based and spirometry-based COPD labels (summarized in Supplementary Table **1**).

To train our ML model, we used binary COPD labels defined based on partial medical-records and self-report data. We defined “medical-record-based” COPD as self-reported COPD or the existence of any COPD-related ICD code in the partial medical records collected by the UK Biobank through linkages to a range of primary care, hereafter “GP dataset”, and hospital inpatient admission records, hereafter “HESIN dataset”.

The medical-record-based COPD labels were derived from three sources: self-reported, hospital inpatient (HESIN) billing codes, and primary care or general practitioner (GP) read codes. Self-reported COPD status was extracted from code 6 (emphysema/chronic bronchitis) of field 6152; and from codes 1112 (chronic obstructive airways disease/copd), 1113 (emphysema/chronic bronchitis), and 1472 (emphysema) of field 20002. COPD cases were identified by the presence of an ICD9 code of 491* (chronic bronchitis), 492* (emphysema), or 496* (chronic airway obstruction) in fields 41271, 41203, or 41205; and by the presence of an ICD10 code of J41* (mucopurulent chronic bronchitis), J42* (unspecified chronic bronchitis), J43* (emphysema), or J44* (other chronic obstructive pulmonary disease) in fields 41270, 41202, or 41204 (Supplementary Table **34**). Cases were also identified by the presence of a v2 or v3 read code in the GP clinical events table (field 42040) corresponding to one of the preceding ICD10 codes via the mappings in fields 1834 (v2) or 1835 (v3). Any individual with evidence of COPD based on at least 1 of the 3 above non-spirometry-based sources was considered a case.

Two sets of patients were defined based on the availability of data from self-report, HESIN, and GP. The *medical-record-based* set (*n* = 325, 027) includes individuals of European ancestry with data available from at least 1 of self-report, HESIN, or GP, whereas the *evaluation* medical-record-based set (*n* = 125, 786) restricts to individuals with data available from all 3 of self-report, HESIN, and GP.

For future hospitalization and death, the date of spirometry assessment was extracted from field 3060 (time of blow measurement). Subsequent hospitalization was identified by the presence of a COPD-related ICD code in the HESIN data (fields 41259 and 41234) dated after the spirometry assessment, and subsequent death was identified by the presence of a COPD-related ICD code in field 40001 (primary cause of death).

Spirometry-based labels mirroring the GOLD criteria (hereafter, proxy GOLD labels) were defined using FEV_1_ and FVC measurements from fields 3062 and 3063. Blows having an extreme value, defined as having any of FEV_1_, FVC, or peak expiratory flow (PEF; field 3064) outside the lower or upper 0.5th percentile, were removed, as were unacceptable blows, identified by having a value other than 0 (no problem) or 32 (user accepted) for field 3061. If multiple valid blows remained, the best ranked blow according to field 3059 was selected, along with the corresponding values of FEV_1_, FVC, and PEF. Sex-specific linear regression models of the following form were developed to predict FEV_1_ on the basis of age and height:

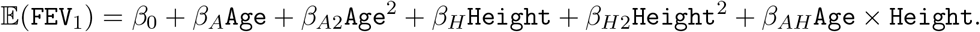

Ancestry was not included as a covariate because subsequent analyses were restricted to individuals of European ancestry. Following the notes for field 20153, the sex-specific FEV_1_ prediction models were trained among healthy never smokers with reproducible spirometry measurements who did not report having wheeze or other respiratory diseases, such as asthma and COPD. A subject was defined as a case with respect to the proxy GOLD 2-4 labels if their FEV_1_/FVC ratio was *<* 0.7 and their observed FEV_1_ was *<* 80% of that predicted for their sex, age, and height.

### 4.3 Machine learning and PRS dataset generation

We generated two datasets for use in the model training and application procedures (Figure **1**). A *modeling* dataset was used to select model architectures, tune hyperparameters, and evaluate ML model performance across tasks while a two-fold *cross-fold* dataset was used during the final model application process to generate phenotypes (Figure **4**). The modeling dataset consisted of training and validation sets containing 80% and 20% of European-ancestry samples with valid spirometry blows, respectively (*n* = 325, 027; Sections 4.1 and 4.2). Samples were randomly assigned to a subset and any individuals with estimated genetic relations (UKB field 22012) spanning these splits were removed to prevent information leakage. The cross-fold dataset further split these training and validation sets into two folds and removed any relations spanning these folds. The random sampling and cross-folding procedures resulted in a similar distribution of labels and spirometry metrics across sets (Supplementary Table **35** and Table **36**). Finally, we defined the *PRS holdout* set to contain the remaining 110,739 European-ancestry individuals with valid genomic information not included in the modeling dataset. By construction, these samples do not have valid blows and were not used in either model training, evaluation, or GWAS.

### 4.4 Machine learning model training and application

We first trained various deep learning models to predict medical-record-based COPD status from spirograms using the *modeling* dataset splits described in section 4.3. We considered a variety of model backbone architectures, including multi-layer perceptrons (MLP), one-dimensional convolutional neural networks (CNNs) [22, 23], and one-dimensional variants of the ResNet9 and ResNet18 networks [24, 25] (Supplementary Table **2**; Supplementary Figure **1** and Figure **2**). Networks were optimized in an end-to-end manner using Adam algorithm [50] to minimize the training cross-entropy loss between the model’s predicted probabilities and the binary COPD status labels. Models were trained for at most 1,500 epochs. In order to prevent overfitting, we employed an early stopping [51] patience of 50 epochs and selected only the checkpoint that resulted in the minimum validation loss. All models were implemented using TensorFlow 2.0 [52] and each model instance was trained on a single NVIDIA Tesla V100 GPU using mixed floating-point precision [53].

For each architecture, we performed a large scale hyperparameter sweep using the Vizier opti mization service [54] (Supplementary Table **37**). Utilizing a Gaussian process bandit optimization algorithm [55] to select hyperparameters for each subsequent run, Vizier ran a total of 150 trials with at most 50 trials running in parallel. For each architecture, we trained ten separate networks using the set of hyperparameters that minimized validation loss in the Vizier [54] sweep (Supplementary Table **38**). Network predictions were averaged to form a mean-ensemble of ten members [56]. Each member was trained using a different seed to ensure random weight initialization and data shuffling, which has been shown to be sufficient for network diversity [57]. These ensembled predictions were then used to evaluate model performance across the COPD status, future hospitalization, and death tasks (Section 2.2).

Baseline MLP and linear models predicting medical-record-based COPD status from only derived spirometry metrics (FEV_1_, FVC, FEV_1_/FVC ratio, and PEF) were trained in a similar manner (Supplementary Table **2** and Table **38**). In contrast to the full spirogram preprocessing described in Section 4.1, these unstructured scalar-valued inputs were simply normalized and standardized.

We generated model predictions for use in GWAS by retraining candidate models on the two *cross-fold* dataset splits described in Section 4.3 (Figure **3**). Similarly to the ensembling process described above, we selected the set of hyperparameters that minimized validation loss in the Vizier sweep and trained two ensembles, each containing ten members, on both folds. Each cross-fold ensemble is then applied to samples from the other fold, ensuring that all ML-based risk predictions are not from the given ensemble’s training split. We combined the two prediction sets to define the final ML-based COPD phenotype. In order to evaluate the effect of training dataset size on model performance and ensure that this cross-fold process did not significantly degrade evaluation metrics, we performed an ablation studying using the best Vizier hyperparameter configuration for the flow-volume ResNet18 model by randomly subsampling the *modeling* training set split to size *n* ∈ {0.1, 0.2, …, 0.9} where each dataset is a strict subset of all larger datasets. We then trained a single model on each dataset using a fixed random seed and compared model performance across tasks using the full *modeling* dataset validation split (Supplementary Figure **5**).

### 4.5 Genome-wide association studies

GWAS analysis of FEV_1_/FVC, medical-record-based COPD status, and ML-based COPD was performed using BOLT-LMM v2.3.6 [28, 58]. For FEV_1_/FVC and medical-record-based COPD status, the GWAS adjusted for age, sex, height, age *×* sex (i.e., an age and sex interaction), age *×* age, and height *×* height, genotyping array, and the top 15 genetic principal components (PCs) (Supplementary Table **39**). The GWAS for ML-based COPD adjusted for all covariates included in the FEV_1_/FVC GWAS, with the addition of a “model-fold” covariate indicating whether a sample was in the first or the second fold of training. The model-fold indicator was included to evaluate and adjust for potential covariate imbalance between the 2 training and prediction folds.

To minimize confounding, the sample was restricted to subjects of European ancestry. Genotypes were filtered to include only autosomal variants with a minor allele frequency (MAF) ≥ 0.001, an imputation INFO score ≥ 0.8, and a Hardy-Weinberg equilibrium (HWE) ≥ 10^−10^.

BOLT-LMM, which fits a linear mixed-effects model, was also used to analyze the proxy GOLD and medical-record-based COPD, which are binary traits. Because these traits are not rare (prevalence = 4.66%) and the sample is neither highly structured nor sampled in an outcome-dependent manner (as in case-control studies), use of BOLT-LMM is expected to be appropriate. As a sensitivity analysis, we repeated the proxy-GOLD COPD GWAS using Regenie [59] and observed very similar results (Supplementary Figure **19**).

### 4.6 Overall survival analysis

Analysis of overall survival (OS) was performed using the time from spirometry ascertainment (field 3060) to death from any cause (field 40000). Subjects who were not known to have died were right-censored at the date of data ingestion (2018-02-12). The association between OS and ML-based COPD risk was quantified using the hazard ratio, which was estimated from a Cox proportional hazards model adjusting for age and sex. The proportional hazards assumption, with respect to COPD risk, was assessed using the Schoenfeld residual test. After stratifying patients into COPD risk quartiles, the OS curves in Figure **3**g were constructed using the standard Kaplan-Meier estimator with point-wise confidence intervals.

### 4.7 SNP-heritability and genetic correlations estimates using GWAS summary statistics

We utilized stratified LD score regression (S-LDSC) [30, 60] to compute the SNP-heritability and genetic correlations using the 75 baseline LD annotations provided by S-LDSC web-page (see URLs).

### 4.8 Replication of ML-based COPD and existing GWAS hits/loci

Top hits were identified using PLINK’s (see URLs) –clump procedure. Linkage disequilibrium (LD) was calculated using a reference panel of 10,000 randomly sampled unrelated individuals of European ancestry. The span of each hit is defined as the linear extent of reference panel variants in LD with the hits at *R*^2^ ≥ 0.1. *Loci* were defined by merging hits that were separated by 250 or fewer Kbp. Two GWAS *G*_1_ and *G*_2_ were compared the counting the numbers of shared and unique hits.

A hit *H*_1_ ∈ *G*_1_ was classified as shared if its span overlapped with the span of any hit from *G*_2_, otherwise it was considered unique. Note that, because a single hit from *G*_1_ can overlap multiple hits from *G*_2_ and vice versa, GWAS comparison is asymmetric.

We compared our GWAS hits/loci with the GWAS catalog (see URLs) using the same method for comparing two GWAS described above. We used the v1.0.2 associations released in July 2022 and converted coordinates from GRCh38 to GRCh37 using UCSC LiftOver (see URLs) with default parameters. All catalog variants whose “DISEASE/TRAIT” column matched the phenotype of interest and were genome-wide significant were converted into loci by merging variants within 250 Kbp.

### 4.9 Hits Simple PRS

To evaluate the quality of hits for each GWAS, we examined their collective predictive power for detecting high risk COPD cases by combining them into a simple PRS. For each GWAS, we defined simple PRS by adding the effects of the GWAS hits (where hits are defined as in Section 4.8). The effect of a hit is defined on its index variant (i.e., the most significant variant of the hit) by multiplying the number of effect alleles by the effect size of the index variant:

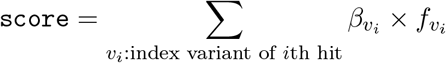

where 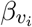 is the effect size from the GWAS summary stats and 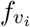 is the number of effect alleles of variant *v*_*i*_.

### 4.10 Tissue/Cell-type specific enrichment analysis of ML-based COPD hits

We utilized two methods to perform tissue/cell-type specific enrichment analysis. First, we utilized the tissues specific analysis in S-LDSC [30, 60] where we utilized 53 baseline version 1 annotations (see URLs), “Multi_tissue_gene_expr” (includes both GTEx [61] and Franke lab data [62, 63]) and “Multi_tissue_chromatin” (includes both Roadmap [64, 65] and EN-TEX data). In the case of gene expression, we utilize 53 tissues or cell types created by Finucane et al.[60] while Franke lab data consists of 152 tissues or cell types. In the case of chromatin data, Roadmap [64, 65] has 397 cell-type- or tissue-specific annotations while EN-TEX data has 93 cell-type- or tissue-specific annotations. As recommended by the S-LDSC authors, we used the -log (p-value) of regression coefficient (*τ*) as the metric to pick the specific tissue or cell-type. Second, we utilize GARFIELD [34] to perform tissue-specific analysis where we utilized 424 DNase I hypersensitive site hotspot annotations provided by the GARFIELD authors [34] and we used the default parameters.

### 4.11 Functional analyses with GREAT

We utilized GREAT v4.0.4 [35] on the human GRCh37 assembly to perform functional enrichment analysis of ML-based COPD risk loci. The default “basal+extension” region-gene association rule was used with 5 kb upstream, 1 kb downstream, 1000 kb extension, and curated regulatory domains included. GREAT analyzes enrichment of terms drawn from multiple data sources including Gene Ontology Biological Process (GOBP), the Mammalian Phenotype Ontology for phenotypes induced by a single gene knockout (MP1KO), and the Human Phenotype Ontology (HP). We considered terms to be statistically significant if the Bonferroni-corrected P-values for both the region-based and gene-based tests were ≤ 0.05.

## Supporting information

Large Supplementary Tables

## Data Availability

Genotypes and phenotypes are available for approved projects through the UK Biobank study. This research has been conducted under Application Number 65275. We utilized the GWAS Catalog for replication analysis. This research used data generated by the COPDGene study (dbGaP accession phs000179.v6.p2), which was supported by NIH grants U01 HL089856 and U01 HL089897. The COPDGene project is also supported by the COPD Foundation through contributions made by an Industry Advisory Board comprised of Pfizer, AstraZeneca, Boehringer Ingelheim, Novartis, and Sunovion. ICGC (International COPD Genetics Consortium) genome-wide association summary statistics were obtained from dbGaP under accession phs000179.v5.p2. SpiroMeta summary statistics were obtained from LDHub.

## URLs

Baseline and BaselineLD annotations: https://data.broadinstitute.org/alkesgroup/ldscore

BOLT-LMM software: https://data.broadinstitute.org/alkesgroup/bolt-lmm

Cell-type specific gene expression annotations: https://alkesgroup.broadinstitute.org/LDSCORE/LDSC_SEG_ldscores/Multi_tissue_gene_expr_1000Gv3_ldscores.tgz

Cell-type specific chromatin annotations: https://alkesgroup.broadinstitute.org/LDSCORE/LDSC_SEG_ldscores/Multi_tissue_chromatin_1000Gv3_ldscores.tgz

GWAS Catalog: https://www.ebi.ac.uk/gwas/

GARFIELD software: https://www.ebi.ac.uk/birney-srv/GARFIELD/

GREAT software: http://great.stanford.edu

PLINK software: https://www.cog-genomics.org/plink1.9

TensorFlow: https://www.tensorflow.org

UCSC LiftOver: https://genome.ucsc.edu/cgi-bin/hgLiftOver

UK Biobank study: https://www.ukbiobank.ac.uk

LDHub: https://ldsc.broadinstitute.org/ldhub

## Data availability

Genotypes and phenotypes are available for approved projects through the UK Biobank study (https://www.ukbiobank.ac.uk) This research has been conducted under Application Number 65275. We utilized the GWAS Catalog (https://www.ebi.ac.uk/gwas/) for replication analysis. This research used data generated by the COPDGene study (dbGaP accession phs000179.v6.p2), which was supported by NIH grants U01 HL089856 and U01 HL089897. The COPDGene project is also supported by the COPD Foundation through contributions made by an Industry Advisory Board comprised of Pfizer, AstraZeneca, Boehringer Ingelheim, Novartis, and Sunovion. ICGC (International COPD Genetics Consortium) genome-wide association summary statistics was obtained from dbGaP under accession phs000179.v5.p2. SpiroMeta summary statistics was obtained from LDHub.

## Code Availability

Code and detailed instructions for model training, prediction, and analysis, as well as instructions for evaluating the trained model on spirograms, are available at https://github.com/Google-Health/genomics-research/tree/main/ml-based-copd.

## Acknowledgements

B.D.H. is supported by NIH K08 HL136928, U01 HL089856, R01 HL155749, and a Research Grant from the Alpha-1 Foundation. M.H.C. is supported by R01HL153248, R01HL149861, R01HL147148, and R01HL089856. D.H. was supported by NIH 2T32HL007427-41.

