## Supplementary Figures and Tables for "Leveraging deep-learning on raw spirograms to improve genetic understanding and risk scoring of COPD despite noisy labels"

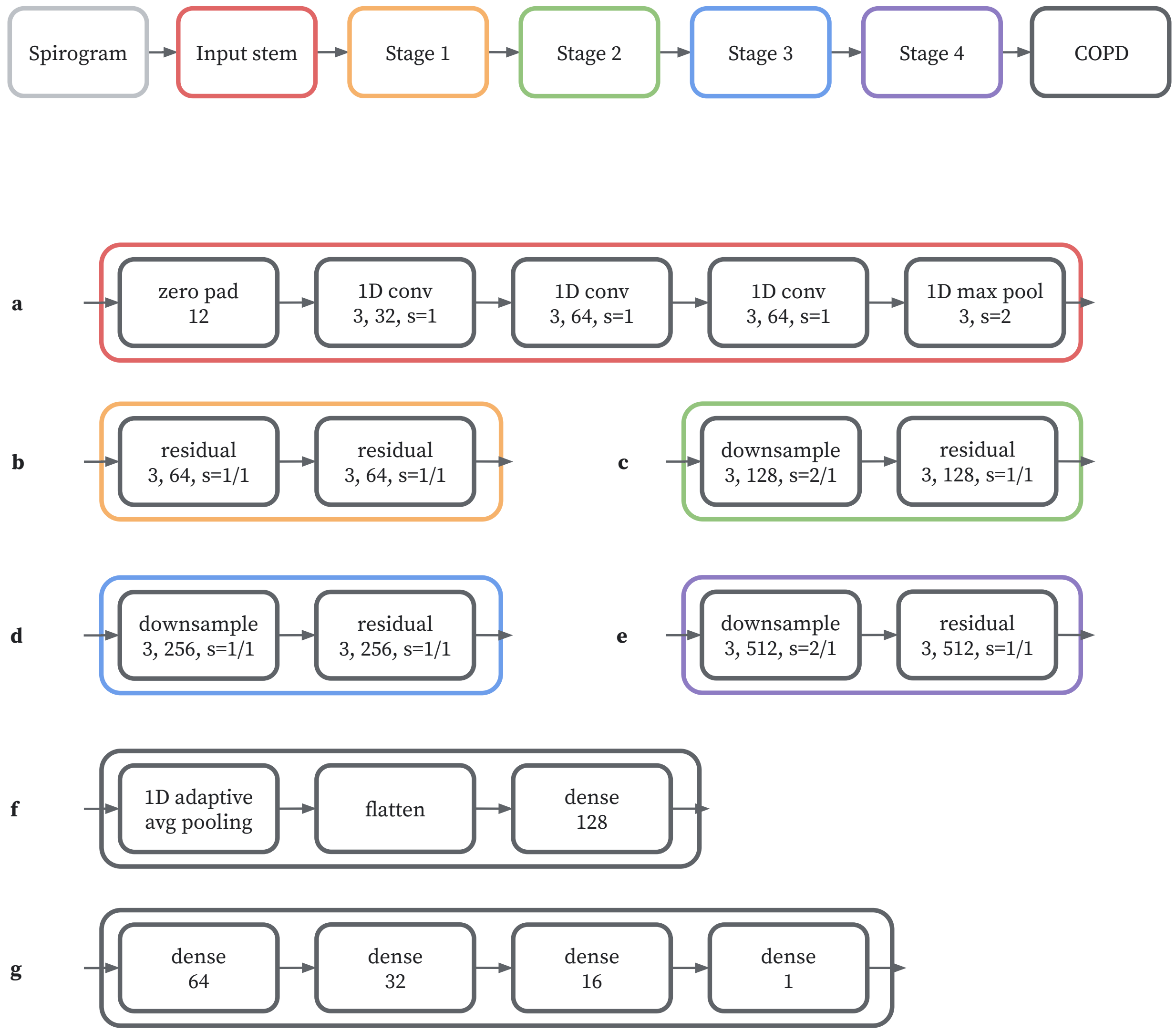


**Supplementary Fig. 1**: An overview of the optimized one-dimensional ResNet18-D model architecture [S24, S25]. Sublabels denote layer parameters, with convolutional, pooling, residual, and downsample layers adhering to the following convention: kernel size, number of filters, and stride length. Supplementary Figure 2 details the residual and downsample layers. Subfigures 1a-f comprise the ResNet18-D model's "backbone", which produces 128-dimensional embeddings from flow-volume spirograms. a) The input stem. We zero-pad the flow-volume curve to size 1024. All 1D convolutions in the input stem are followed by a batch normalization layer [S66] and ReLU [S67] activation function. b-e) Residual and downsampling layers (Supplementary Figure 2). f) The backbone's final layer, followed by a ReLU activation function, produces an 128-dimensional embedding passed to each outcome head. g) The model's disease head subarchitecture. We use swish activation functions [S68] in the first three layers and a sigmoid activation function in the final layer.

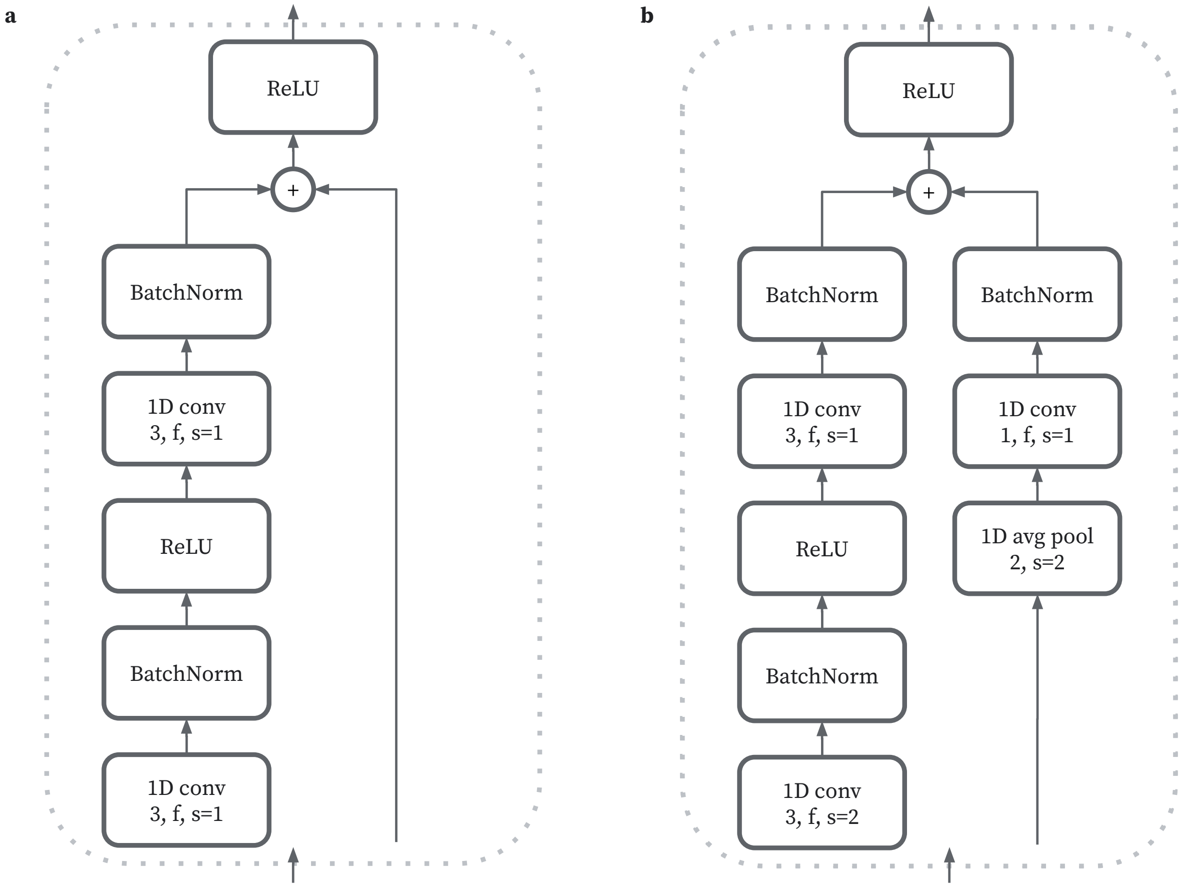


**Supplementary Fig. 2**: An overview of the one-dimension ResNet18-D residual and downsample layers. a) A basic residual layer [S24]. b) A downsampling residual layer following ResNet18-D architecture modifications [S25].

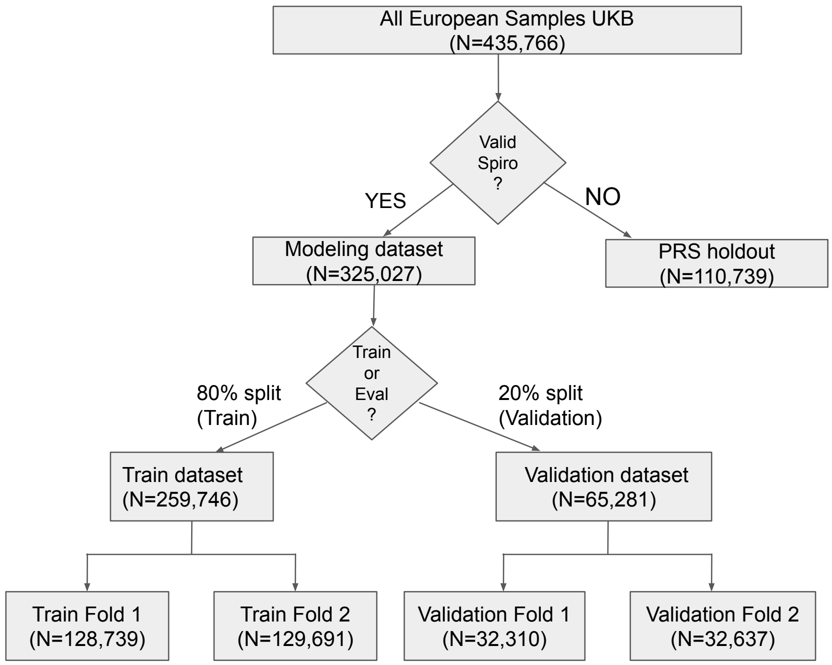


**Supplementary Fig. 3**: An overview of UK Biobank dataset used in this study. Our initial dataset consists of all European-ancestry in UK Biobank ($n = 435,766$). We considered all individuals with valid spirograms as modeling dataset ($n = 325,027$) and individuals with invalid spirograms are used as PRS holdout set. The PRS holdout set is from the European individuals who are not used in the ML modeling and in the GWASs ($n$=110,739). We split the modeling datasets to train and validation set with 80% and 20% of samples, respectively. The *modeling* dataset was used to select model architectures, tune hyperparameters, and evaluate ML model performance across tasks while a two-fold *cross-fold* dataset was used during the final model application process to generate phenotypes. It is worth mentioning that the combination of train fold 1 and train fold 2 sample size is not equal to the size of the whole training dataset due to the fact we removed genetically close samples that fall cross two different folds. As folds were constructed to keep genetically related individuals together, preventing the same individual or a close relative from being used for both training and prediction.

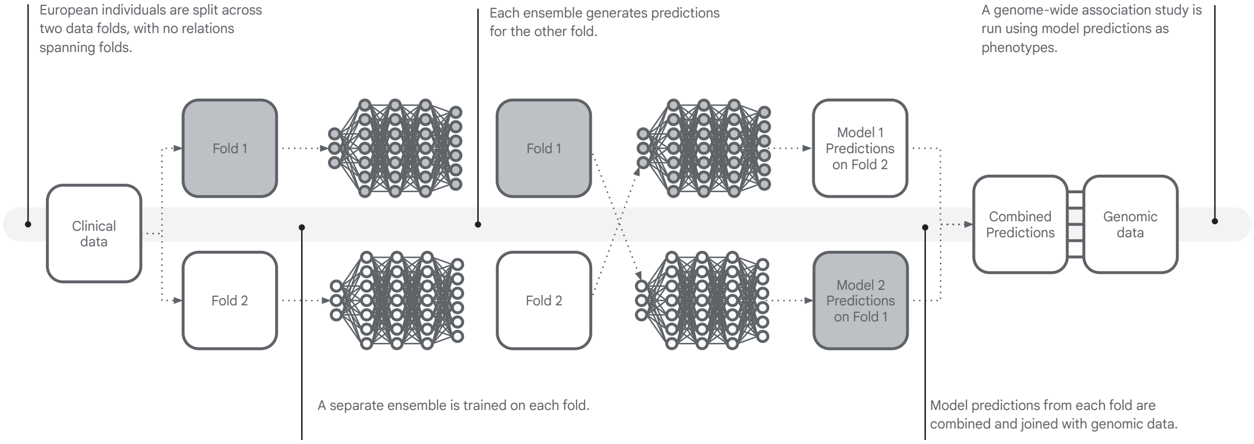


**Supplementary Fig. 4**: An overview of the cross-fold training and application process

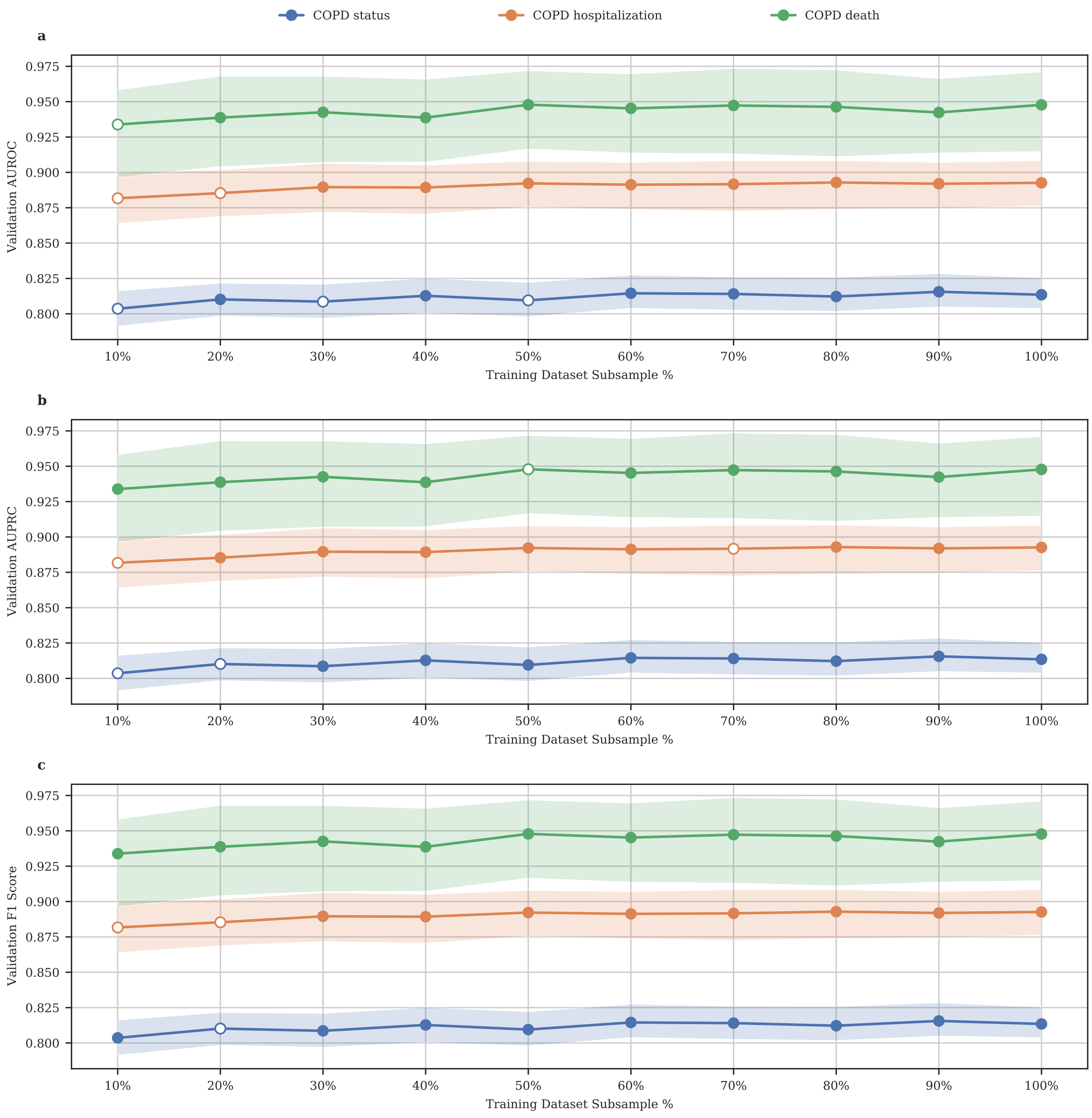


**Supplementary Fig. 5**: An ablation study assessing the impact of training dataset size on ResNet18 model performance. Ten separate ResNet18 models were trained on subsampled versions of the full training dataset (n=259,748) and then evaluated across the COPD status, hospitalization, and death tasks using the full validation dataset. Each subsampled dataset is a valid subset of all larger datasets. For example, all samples in the 40% dataset are also contained in the 50-100% datasets. Each point in the figure denotes the average metric over 100 bootstrapping samples while error bars denote the associated 95% confidence intervals. Unfilled points represent metrics with statistically significant differences compared to the corresponding 100% model under paired boostrapping. a) A comparison of validation AUROC across dataset sizes. b) A comparison of validation AUPRC across dataset sizes. c) A comparison of validation F1 scores across dataset sizes.

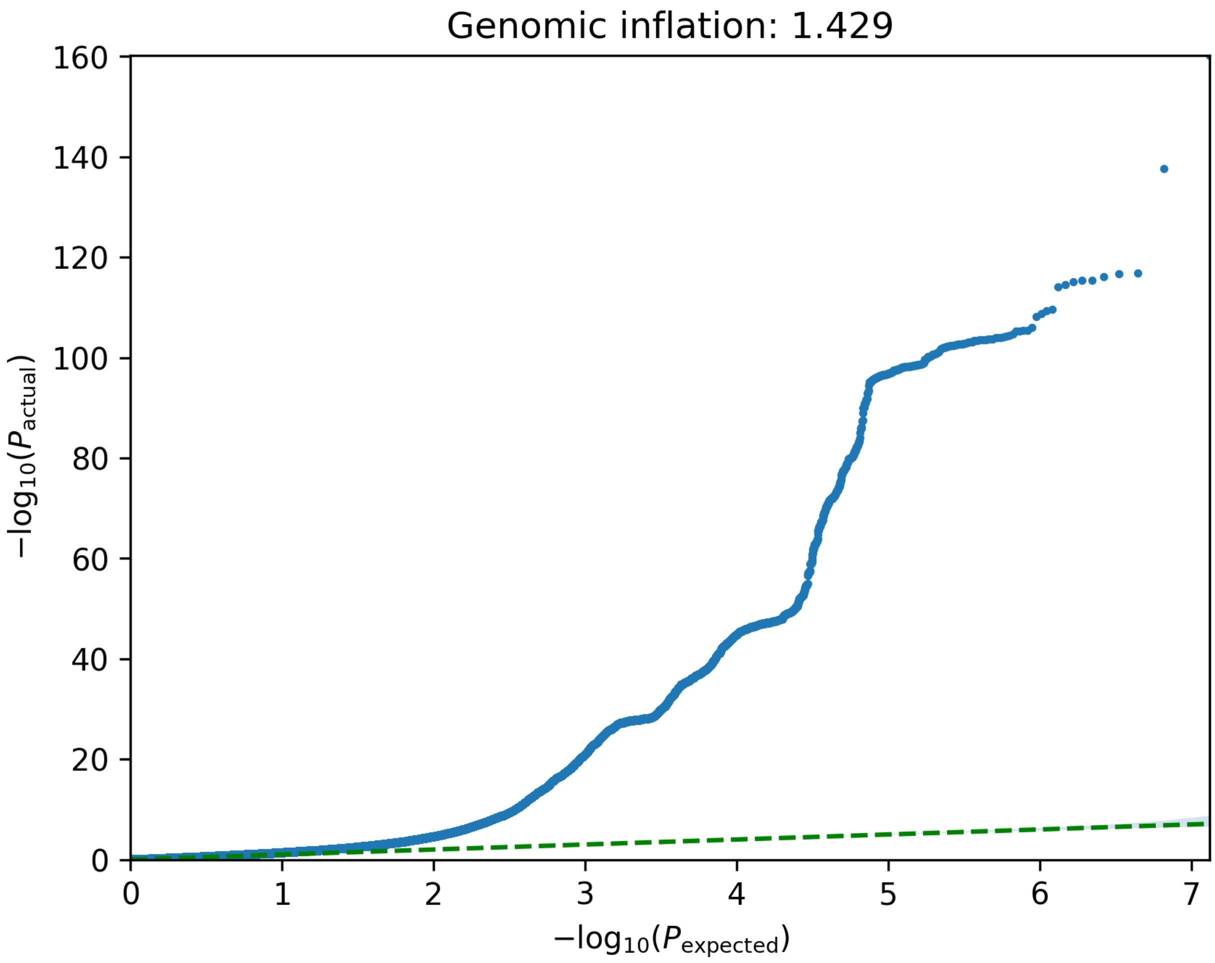


**Supplementary Fig. 6**: **QQplot of ML-based COPD GWAS.**

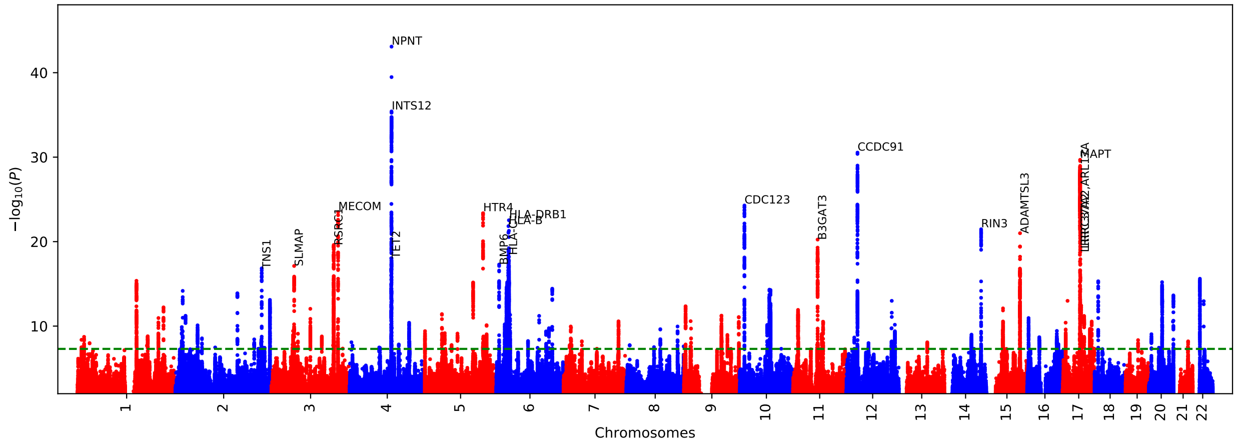


**Supplementary Fig. 7**: **ML-based COPD GWAS Manhattan plot conditional on $FEV_1$/FVC.**

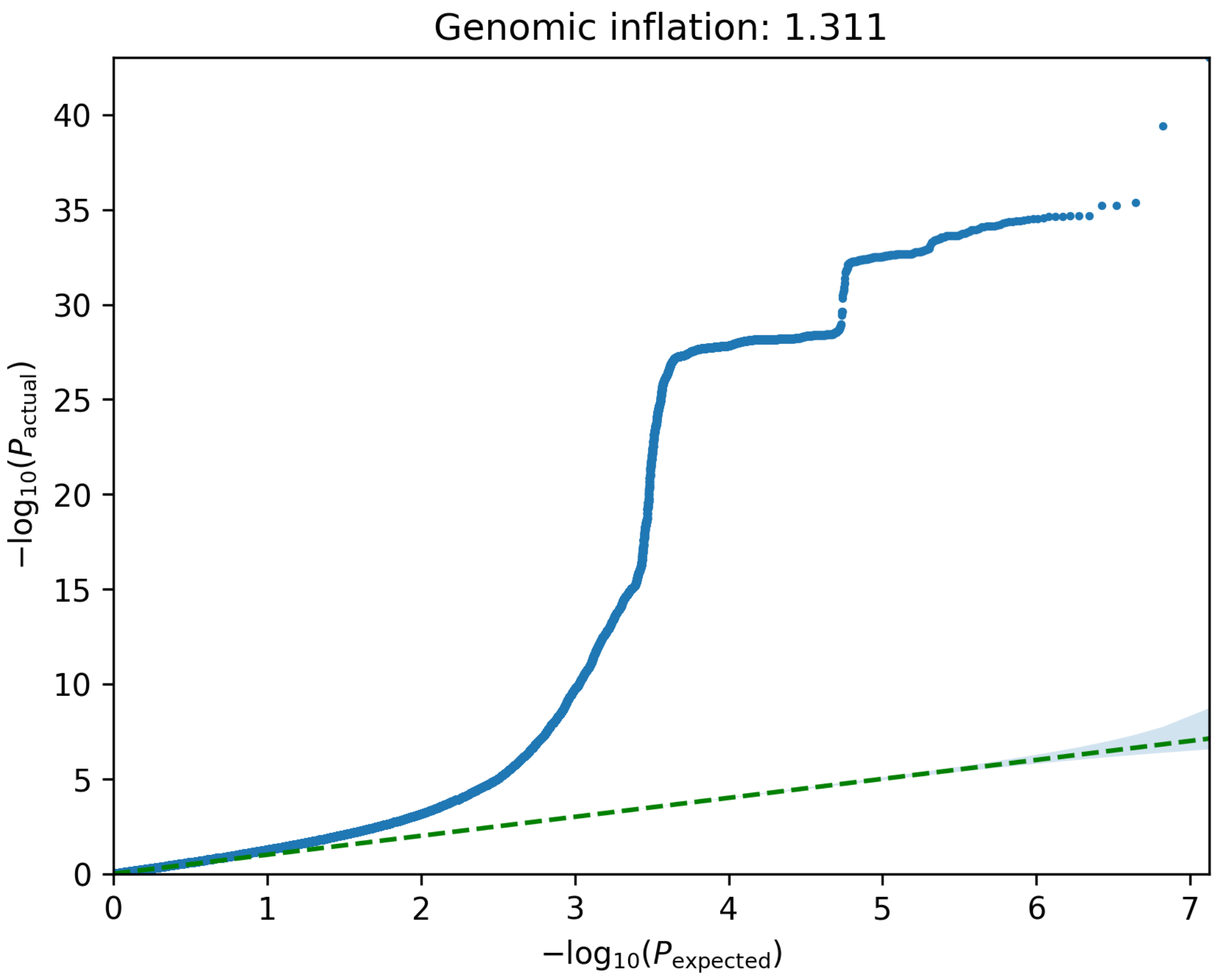


**Supplementary Fig. 8**: **QQplot of ML-based COPD GWAS conditional on $FEV_1$/FVC.**

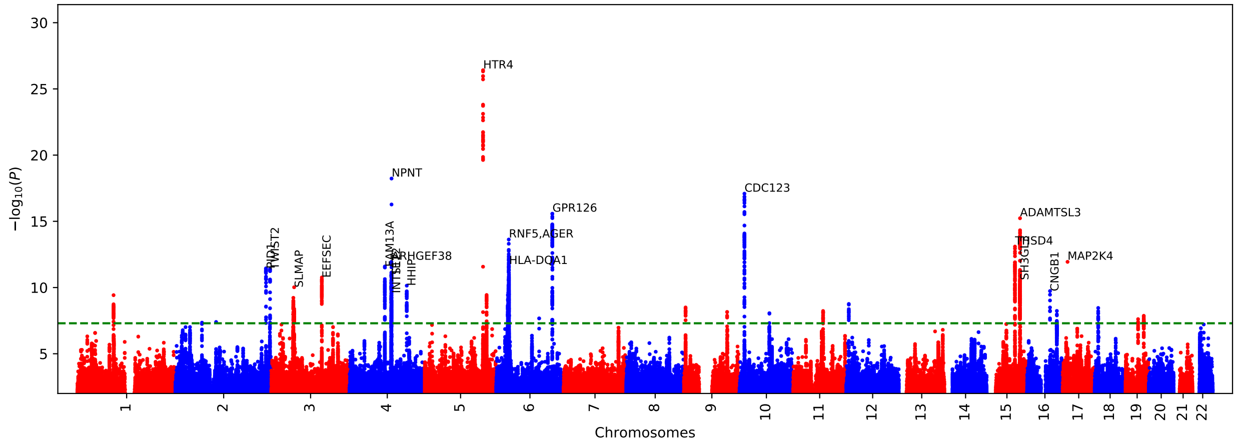


**Supplementary Fig. 9**: **ML-based COPD GWAS Manhattan plot conditional on $FEV_1$/FVC, $FEV_1$, FVC, and PEF.**

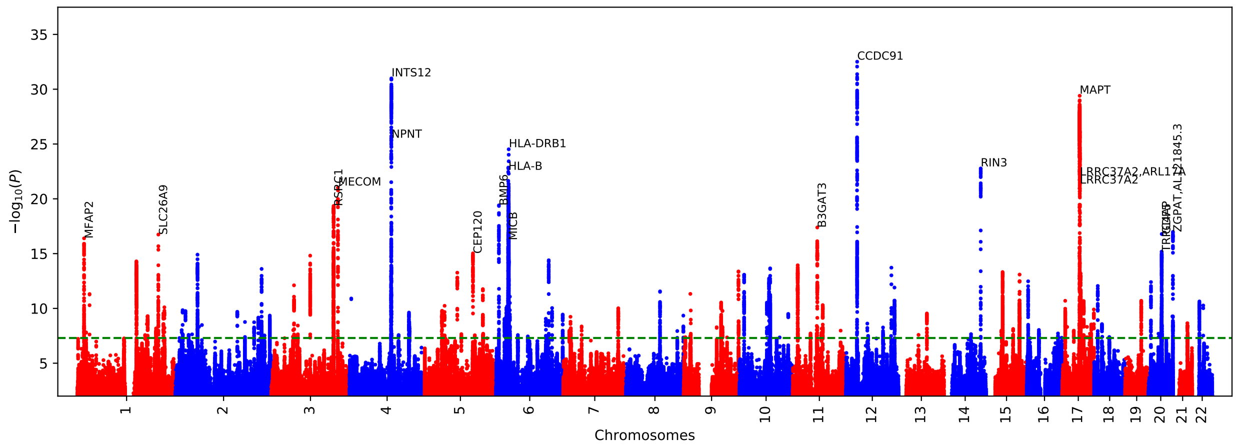

**Supplementary Fig. 10**: **ML-based COPD GWAS Manhattan plot via DeepNull.** We performed ML-based COPD GWAS where we used the same set of covariates as the Figure 4 with one additional covariate provided by DeepNull [S31]. DeepNull model predicts the ML-based COPD using age, sex, genotype-array, and $FEV_1/FVC$ as inputs. The additional DeepNull-covariate is the DeepNull model prediction of ML-based COPD. DeepNull learns a function (i.e., linear or non-linear) that predicts ML-based COPD via age, sex, genotype-array, and $FEV_1/FVC$ as inputs. Thus, this analysis is similar to the ML-based COPD GWAS conditional on $FEV_1/FVC$ where instead of assuming that $FEV_1/FVC$ has linear relationship with ML-based COPD, DeepNull handles cases where age, sex, and $FEV_1/FVC$ can have non-linear relationship with ML-based COPD.

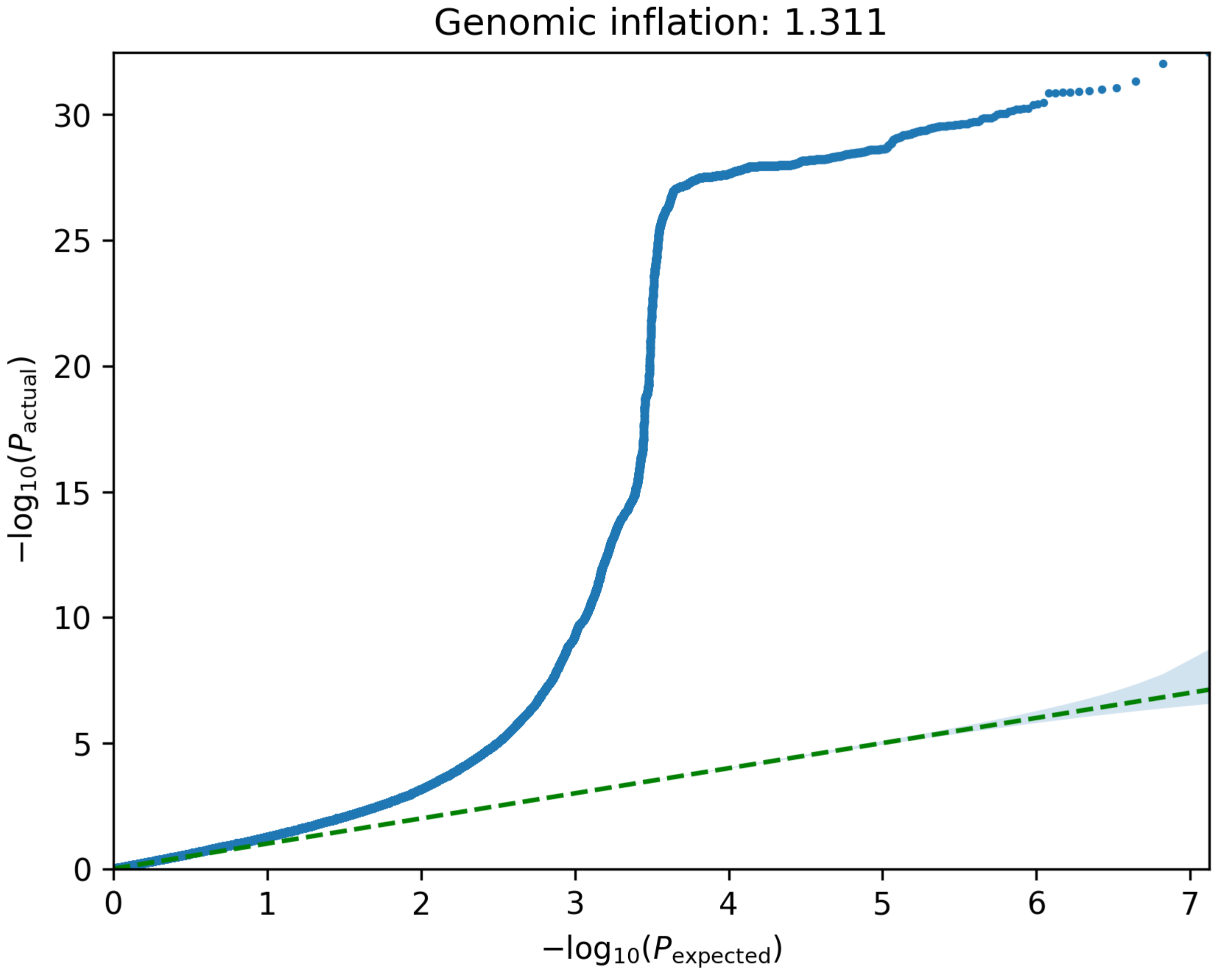


**Supplementary Fig. 11: QQplot of ML-based COPD GWAS via DeepNull.**

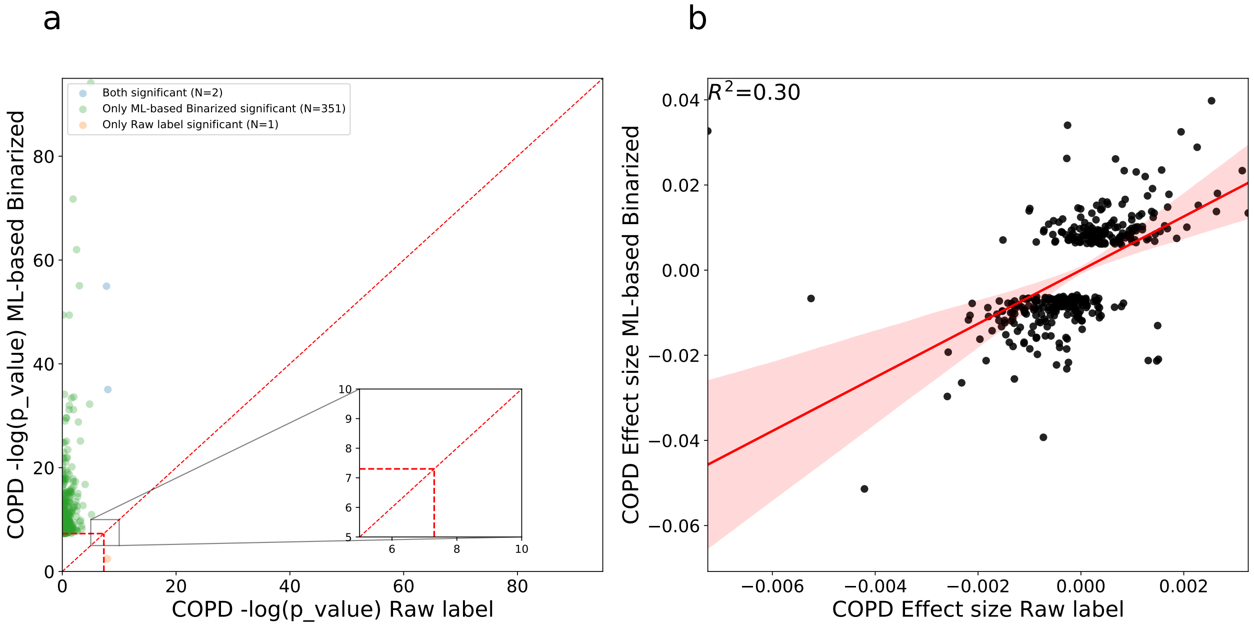


**Supplementary Fig. 12**: **Statistical power comparison of binarized ML-based COPD with medical-record-based COPD labels.** a) The X-axis is the -log p-value of medical-record-based COPD. The Y-axis is -log p-value of the binarized ML-based COPD. Both p-values are computed using two-sided tests. The vertical and horizontal red line indicates the genome-wide significance level. The diagonal red line indicates the y=x. The orange dots indicate variants that are significant for medical-record-based COPD but not significant for our binarized ML-based COPD and green dots indicate variants that are significant for our binarized ML-based COPD but not significant for the medical-record-based COPD label GWAS. b) Effect size correlation of binarized ML-based COPD and medical-record-based COPD GWAS. The X-axis is the effect size of medical-record-based label COPD for all GWS hits and Y-axis is the effect size of our binarized ML-based COPD.

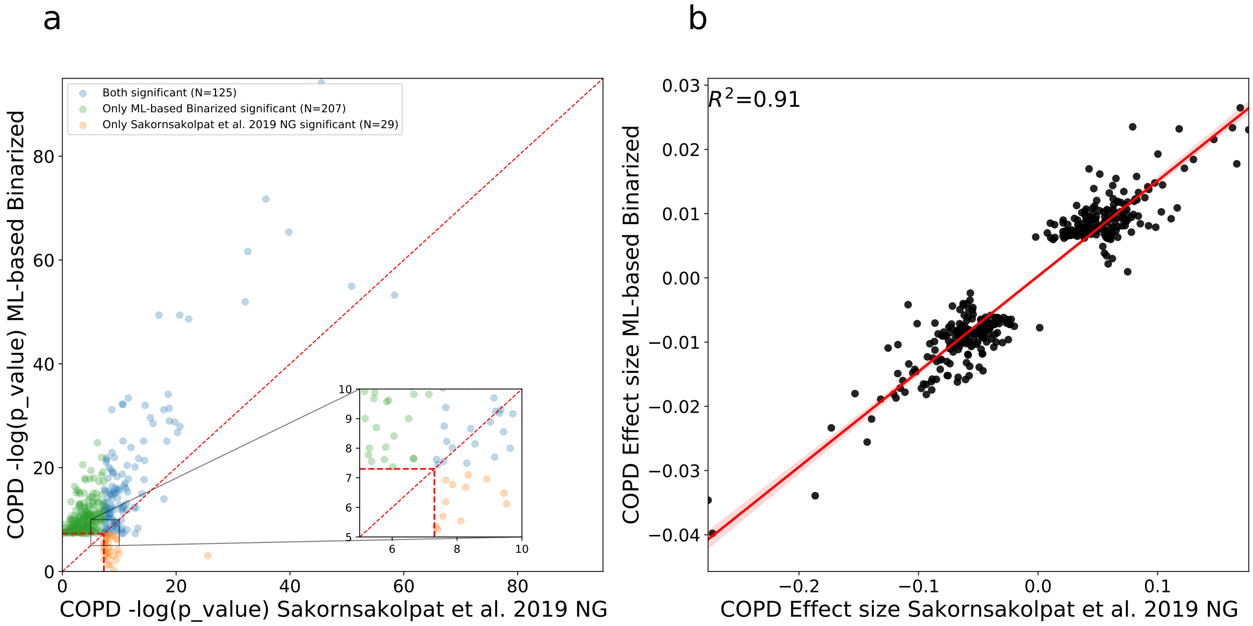


**Supplementary Fig. 13**: **Statistical power comparison of binarized ML-based COPD with Sakornsakolpat.** a) The X-axis is the -log p-value of Sakornsakolpat et al. [S12]. The Y-axis is -log p-value of the binarized ML-based COPD. Both p-values are computed using two-sided tests. The vertical and horizontal red line indicates the genome-wide significance level. The diagonal red line indicates the y=x. The orange dots indicate variants that are significant for Sakornsakolpat et al. [S12] but not significant for our binarized ML-based COPD and green dots indicate variants that are significant for our binarized ML-based COPD but not significant for Sakornsakolpat et al. [S12] GWAS. b) Effect size correlation of binarized ML-based COPD and Sakornsakolpat et al. [S12] GWAS. The X-axis is the effect size of Sakornsakolpat et al. [S12] for all GWS hits and Y-axis is the effect size of our binarized ML-based COPD.

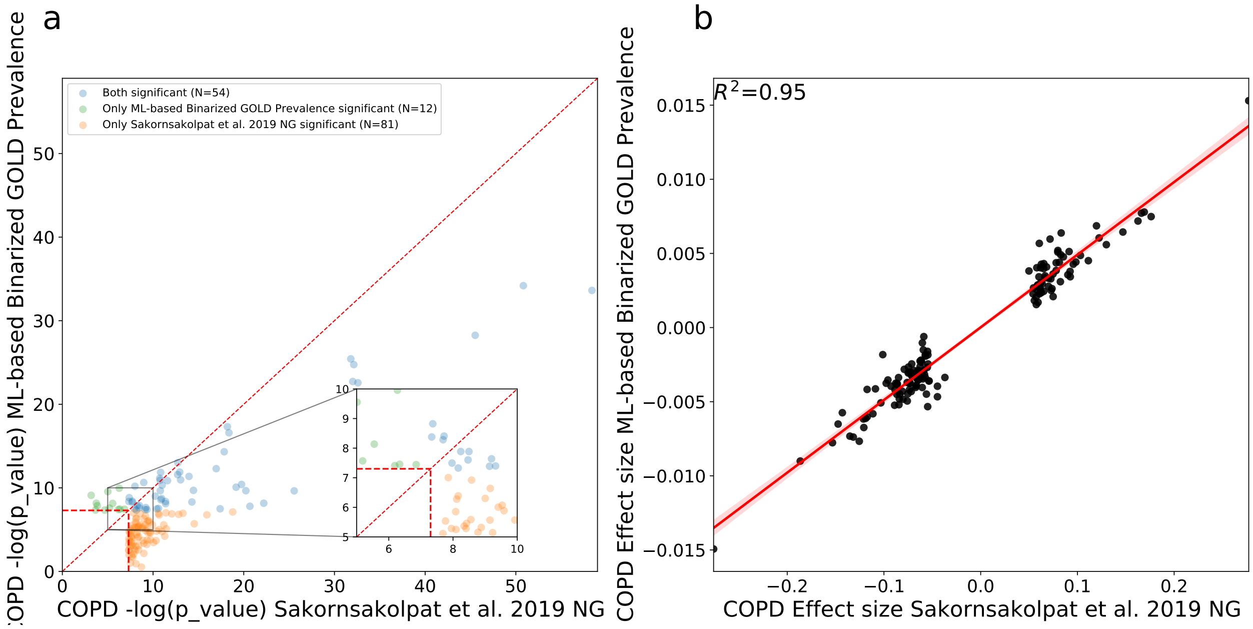


**Supplementary Fig. 14**: **Statistical power comparison of binarized ML-based COPD match GOLD prevalence with Sakornsakolpat et al.** a) The X-axis is the -log p-value of proxy-GOLD. The Y-axis is -log p-value of the binarized ML-based COPD prevalence match with proxy-GOLD. Both p-values are computed using two-sided tests. Both p-values are computed using two-sided tests. The vertical and horizontal red line indicates the genome-wide significance level. The diagonal red line indicates the y=x. The orange dots indicate variants that are significant for Sakornsakolpat but not significant for our binarized ML-based COPD and green dots indicate variants that are significant for our binarized ML-based COPD but not significant for Sakornsakolpat GWAS. b) Effect size correlation of binarized ML-based COPD and Sakornsakolpat GWAS. The X-axis is the effect size of Sakornsakolpat for all GWS hits and Y-axis is the effect size of our binarized ML-based COPD.

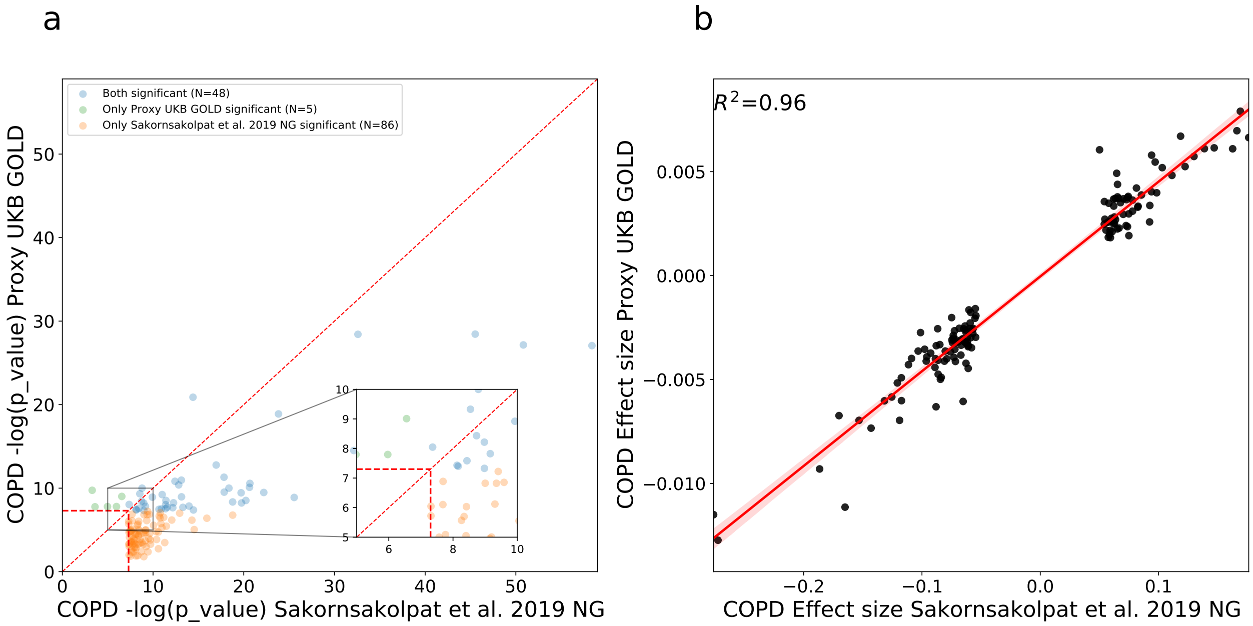


**Supplementary Fig. 15**: **Statistical power comparison of proxy-GOLD with Sakornsakolpat et al.** a) The X-axis is the -log p-value of Sakornsakolpat et al. [S12]. The Y-axis is the -log p-value of proxy-GOLD. The vertical and horizontal red line indicates the genome-wide significance level. The diagonal red line indicates the y=x. The orange dots indicate variants that are significant for Sakornsakolpat but not significant for proxy-GOLD and green dots indicate variants that are significant for proxy-GOLD but not significant for Sakornsakolpat GWAS. b) Effect size correlation of proxy-GOLD and Sakornsakolpat GWAS. The X-axis is the effect size of Sakornsakolpat for all GWS hits and Y-axis is the effect size of proxy-GOLD.

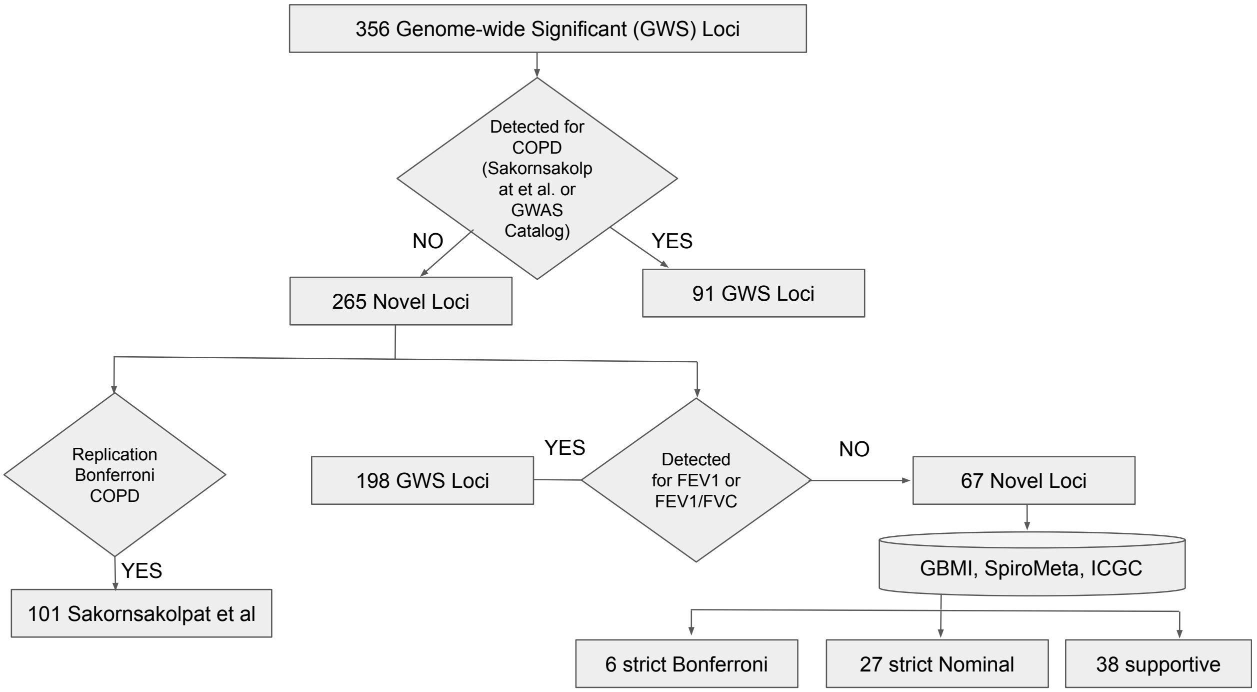


**Supplementary Fig. 16**: A GWAS on the ML-based liability score identifies 265 novel COPD risk loci in addition to 91 previously known COPD loci with respect to [S12] and GWAS catalog entries (as of 2022-07-09) for COPD, emphysema, chronic bronchitis. Out of 265 novel loci, 221 of which independently replicate as associated with COPD or COPD-related lung function as follows. We observed that 101 out of 265 replicate in a previous COPD GWAS [S12] after Bonferroni correction. Also, 198 out of 265 are previously known $FEV_1$ or $FEV_1/FVC$ loci with respect to [S14] and GWAS catalog entries. The three datasets are GBMI (Global Biobank Meta-analysis Initiative) [S32], SpiroMeta [S33], and ICGC (International COPD Genetics Consortium) [S11]. We defined two replication strategies: First, we defined *supportive* replication as consistent effect size direction across all studies with our ML-based COPD. The ICGC and GBMI GWAS are based on a COPD phenotype; thus, we expect their effect size signs to match our ML-based COPD. SpiroMeta phenotypes, on the other hand, capture lung function, so we expect their effect size signs to be the opposite of our ML-based COPD signs. Second, we defined *strict* replication as consistent effect size direction in any study with Bonferroni correction of $P < 0.1$ (one-sided) for that study.

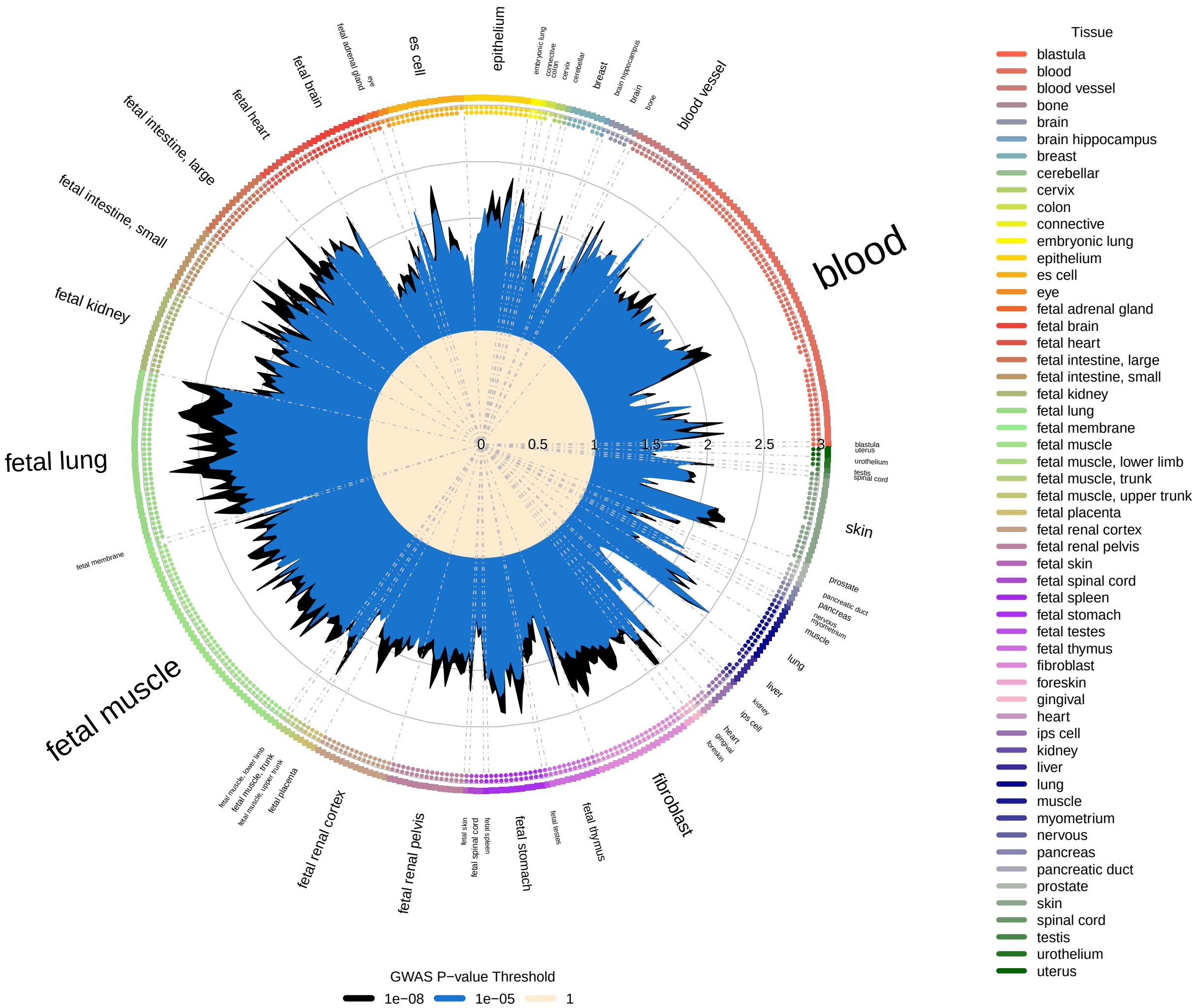


**Supplementary Fig. 17**: **Enrichment overlap of ML-based COPD GWAS with DNase I hotspots computed using GARFIELD.** Radial plot illustrates the enrichment (OR) in each cell type for different GWAS p-value thresholds ($P < 10^{-8}$ and $10^{-5}$). In addition, the small dots on the outer side of the plot indicates enrichment significant level computed by GARFIELD for different significant level of $10^{-5}$, $10^{-6}$, $10^{-7}$, and $10^{-8}$ in direction of outside to insider of plot.

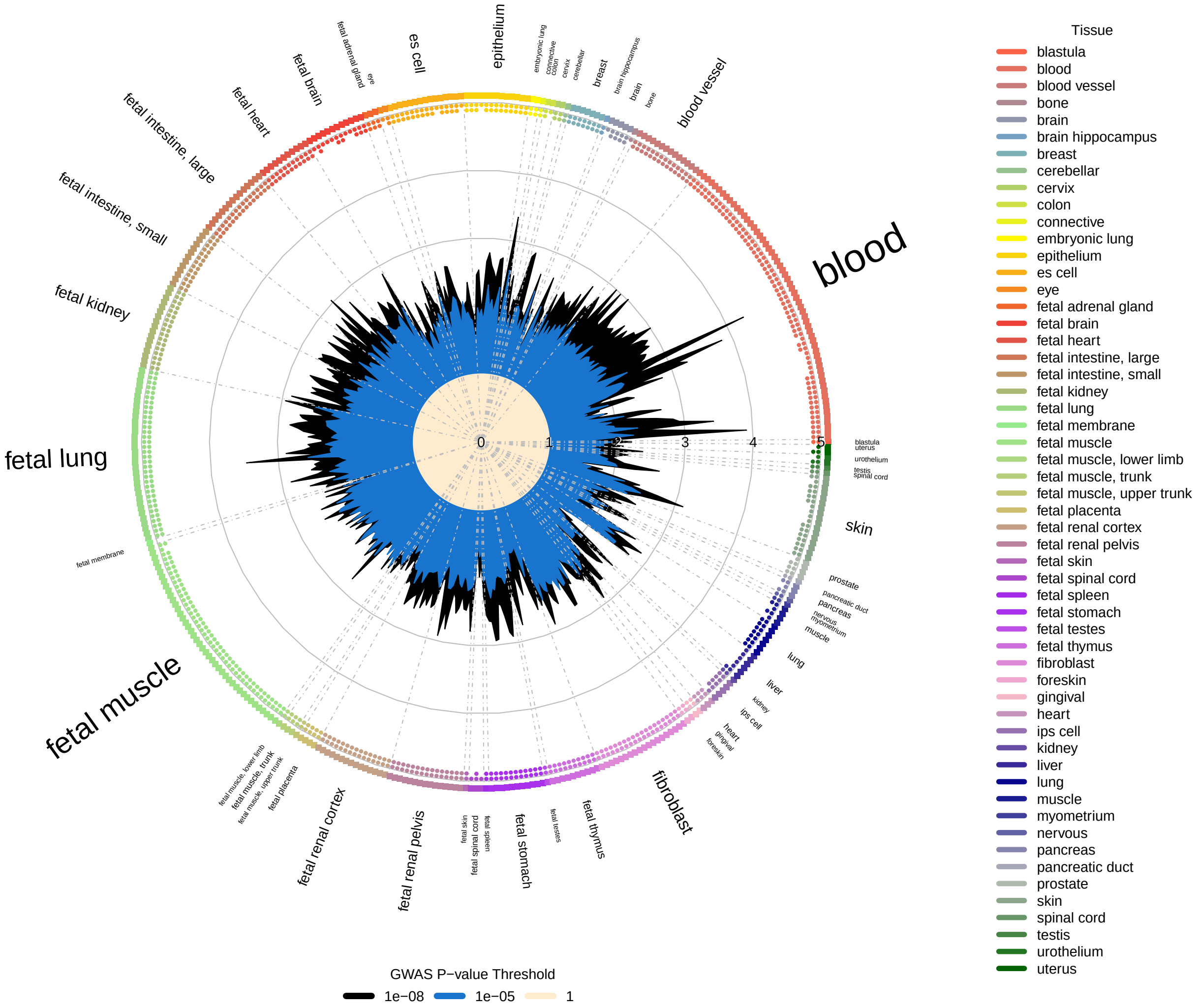


**Supplementary Fig. 18: Enrichment overlap of ML-based COPD conditional on $FEV_1$/FVC GWAS with DNase I hotspots computed using GARFIELD.** We observed the strongest enrichment in blood, fetal lung, and embryonic lung.

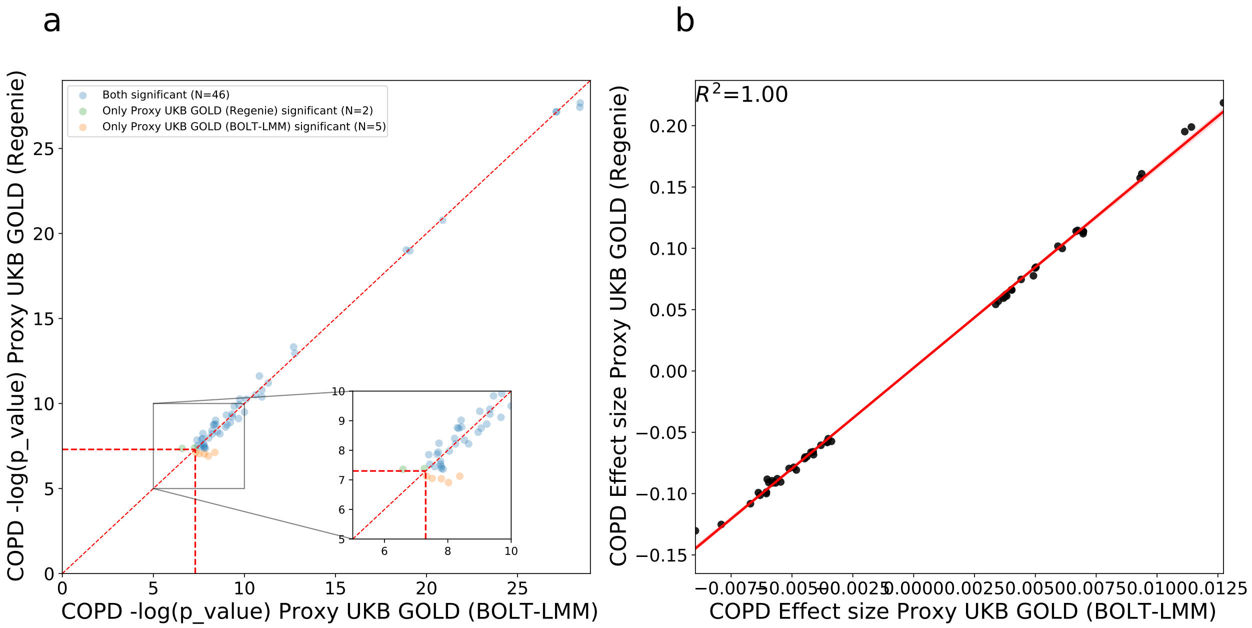


**Supplementary Fig. 19**: **Statistical power comparison of proxy-GOLD label using BOLT-LMM vs Regenie.** a) The X-axis is the -log p-value of proxy-GOLD via BOLT-LMM. The Y-axis is -log p-value of proxy-GOLD via Regenie. Both p-values are computed using two-sided tests. The vertical and horizontal red line indicates the genome-wide significance level. The diagonal red line indicates the y=x. The orange dots indicate variants that are significant for BOLT-LMM but not significant for Regenie and green dots indicate variants that are significant for Regenie but not significant for BOLT-LMM. b) Effect size correlation of proxy-GOLD GWAS via BOLT-LMM and Regenie. The X-axis is the effect size of BOLT-LMM for all GWS hits and Y-axis is the effect size of Regenie obtained from proxy-GOLD COPD.

### Supplementary Tables

| Label | Definition | Usage |
|---|---|---|
| Self report | Code 6 in field 6152 (medical conditions in touch screen questionnaires) or codes 1112, 1113, or 1472 in field 20002 (medical conditions in verbal interview). | Definition of medical-record-based COPD labels below used in training. |
| Primary Care | ICD-10 codes J41, J42, J43, or J44 in field 42040 (GP clinical event records), after Read v2 and v3 codes in the records mapped into ICD-10 codes. | Definition of medical-record-based COPD labels below used in training. |
| Training Hospitalization | Includes primary or secondary causes of hospitalization. ICD-9 codes 491, 492, or 496 in field 41271 (Diagnoses - ICD9), or ICD-10 codes J41, J42, J43, or J44 in field 41270 (Diagnoses - ICD10). | Definition of medical-record-based COPD labels below used in training. |
| Medical-record-based | If a COPD case in at least one of "self report", "primary care", and "training hospitalization" COPD labels. | Training of ML models in Sections 2.1 and 2.2. |
| Evaluation medical-record-based | Logical OR of "self report", "primary care", and "hospitalization" labels only when all three sources exist for an individual. | Evaluation of ML models in Sections 2.2 and 2.3, and GWAS hits in Section 2.7. Having all three sources increases the likelihood of a correct COPD label which is preferred for evaluation. |
| Future hospitalization | Only includes cases with COPD as primary cause of hospitalization *after the spirometry test date*. ICD-10 codes J41, J42, J43, or J44 in field 41234 (records in HES inpatient diagnoses dataset) after converting ICD-9 codes to ICD-10 codes. | Evaluation of ML models in Sections 2.2 and 2.3. |
| Death | ICD-10 J41, J42, J43, or J44 codes in field 40001 (primary cause of death). | Evaluation of ML models in Sections 2.2 and 2.3, and GWAS hits in Section 2.7. |
| Hospitalization | Similar to "future hospitalization" but also includes cases before the spirometry test date. | Evaluation of GWAS hits in Section 2.7. |
| Proxy GOLD | Mirrors moderate or worse GOLD grading for *a single blow* without bronchodilation: $FEV_1/FVC < 0.7$ and $FEV_1$%predicted $< 80\%$. | Evaluating the noisiness of training labels, "medical-record-based" COPD. Evaluation of binarized ML-based COPD liability in Section 2.1. |

**Supplementary Table 1**: **Definition of the binary COPD labels in UKB.** These labels were used for both training and evaluating ML models as well as running label-based GWAS.

See the attached Excel table.

**Supplementary Table 2**: **Overview of various model architectures.**

| Model | Medical-record-based AUC | Hospitalization AUC | Death AUC |
|---|---|---|---|
| GOLD 2-4 | 0.6892 (0.6788–0.6991) | 0.7607 (0.7350–0.7802) | 0.8374 (0.7708–0.8978) |
| $FEV_1/FVC$ Ratio | 0.7785 (0.7652–0.7917) | 0.8362 (0.8191–0.8566) | 0.8871 (0.8047–0.9398) |
| $FEV_1$%predicted | 0.7793 (0.7669–0.7909) | 0.8712 (0.8517–0.8878) | 0.9166 (0.8571–0.9654) |
| Spiro-metric LogReg | 0.7976 (0.7852–0.8096) | 0.8748 (0.8545–0.8924) | 0.9348 (0.8981–0.9611) |
| Spiro-metric MLP | 0.8023 (0.7903–0.8142) | 0.8776 (0.8575–0.8958) | 0.9379 (0.9086–0.9628) |
| Flow-volume MLP | 0.8031 (0.7885–0.8146) | 0.8808 (0.8604–0.9006) | 0.9453 (0.9139–0.9720) |
| Flow-volume ResNet18 | 0.8161 (0.8055–0.8295) | 0.8949 (0.8779–0.9106) | 0.9490 (0.9207–0.9713) |

| Model | Medical-record-based AUPRC | Hospitalization AUPRC | Death AUPRC |
|---|---|---|---|
| GOLD 2-4 | 0.1447 (0.1328–0.1562) | 0.0359 (0.0305–0.0410) | 0.0053 (0.0034–0.0075) |
| $FEV_1/FVC$ Ratio | 0.2199 (0.1979–0.2364) | 0.0833 (0.0663–0.1043) | 0.0219 (0.0111–0.0374) |
| $FEV_1$%predicted | 0.2100 (0.1888–0.2309) | 0.0940 (0.0740–0.1144) | 0.0292 (0.0147–0.0512) |
| Spiro-metric LogReg | 0.2546 (0.2305–0.2768) | 0.1078 (0.0862–0.1345) | 0.0288 (0.0157–0.0508) |
| Spiro-metric MLP | 0.2859 (0.2622–0.3112) | 0.1343 (0.1042–0.1662) | 0.0383 (0.0164–0.0774) |
| Flow-volume MLP | 0.3160 (0.2896–0.3389) | 0.1639 (0.1298–0.1976) | 0.0426 (0.0193–0.0820) |
| Flow-volume ResNet18 | 0.3282 (0.3004–0.3527) | 0.1777 (0.1403–0.2100) | 0.0586 (0.0260–0.1223) |

| Model | Medical-record-based F1 | Hospitalization F1 | Death F1 |
|---|---|---|---|
| GOLD 2-4 | - | - | - |
| $FEV_1/FVC$ Ratio | 0.3067 (0.2892–0.3224) | 0.1918 (0.1649–0.2170) | 0.0490 (0.0046–0.0900) |
| $FEV_1$%predicted | 0.2984 (0.2778–0.3199) | 0.1901 (0.1583–0.2201) | 0.0825 (0.0222–0.1544) |
| Spiro-metric LogReg | 0.3435 (0.3217–0.3599) | 0.2100 (0.1796–0.2352) | 0.0567 (0.0136–0.1225) |
| Spiro-metric MLP | 0.3536 (0.3291–0.3739) | 0.2263 (0.1920–0.2556) | 0.1158 (0.0425–0.2046) |
| Flow-volume MLP | 0.3633 (0.3393–0.3852) | 0.2603 (0.2191–0.2967) | 0.1174 (0.0553–0.2151) |
| Flow-volume ResNet18 | 0.3793 (0.3565–0.3993) | 0.2709 (0.2369–0.3040) | 0.1205 (0.0374–0.2253) |

**Supplementary Table 3**: **Comparison of model AUC, AUPRC, and F1 scores across evaluation medical-record-based COPD disease status, future COPD-related hospitalization, and COPD-related death.** GOLD 2-4, $FEV_1/FVC$ Ratio, and $FEV_1$%predicted denote risk models based on standard spirometry metrics. The spirometry metric logistic regression and MLP models were trained to predict COPD status from only $FEV_1/FVC$ ratio, $FEV_1$, FVC, and PEF while the flow-volume MLP and ResNet18 models utilized the entire flow-volume curve. We performed hyperparameter sweeps for each class of deep learning model, selecting the model that minimized the binary cross entropy loss over the modeling validation set. 95% confidential intervals were generated using bootstrapping ($n = 100$ bootstrapping trials).

| Model | Medical-record-based AUC | Hospitalization AUC | Death AUC |
|---|---|---|---|
| GOLD 2-4 | 0.1269 (0.1156–0.1371) | 0.1342 (0.1185–0.1527) | 0.1116 (0.0596–0.1577) |
| $FEV_1$/FVC Ratio | 0.0376 (0.0293–0.0460) | 0.0587 (0.0446–0.0733) | 0.0619 (0.0174–0.1177) |
| $FEV_1$%predicted | 0.0368 (0.0276–0.0460) | 0.0238 (0.0128–0.0398) | 0.0324 (-0.0007–0.0768) |
| Spiro-metric LogReg | 0.0185 (0.0112–0.0245) | 0.0202 (0.0124–0.0277) | 0.0141 (0.0017–0.0285) |
| Spiro-metric MLP | 0.0138 (0.0075–0.0179) | 0.0173 (0.0086–0.0258) | 0.0110 (0.0015–0.0205) |
| Flow-volume MLP | 0.0130 (0.0073–0.0185) | 0.0142 (0.0078–0.0232) | 0.0036 (-0.0048–0.0117) |

| Model | Medical-record-based AUPRC | Hospitalization AUPRC | Death AUPRC |
|---|---|---|---|
| GOLD 2-4 | 0.1836 (0.1636–0.2048) | 0.1418 (0.1077–0.1711) | 0.0533 (0.0212–0.1155) |
| $FEV_1$/FVC Ratio | 0.1084 (0.0912–0.1252) | 0.0944 (0.0704–0.1206) | 0.0367 (0.0122–0.0823) |
| $FEV_1$%predicted | 0.1182 (0.0940–0.1380) | 0.0837 (0.0569–0.1096) | 0.0294 (0.0043–0.0744) |
| Spiro-metric LogReg | 0.0737 (0.0588–0.0914) | 0.0699 (0.0504–0.0912) | 0.0298 (0.0076-0.0738) |
| Spiro-metric MLP | 0.0424 (0.0304–0.0548) | 0.0434 (0.0275–0.0589) | 0.0203 (-0.0003–0.0537) |
| Flow-volume MLP | 0.0122 (0.0031–0.0193) | 0.0138 (0.0020–0.0275) | 0.0160 (0.0004–0.0389) |

| Model | Medical-record-based F1 | Hospitalization F1 | Death F1 |
|---|---|---|---|
| GOLD 2-4 | - | - | - |
| $FEV_1$/FVC Ratio | 0.0725 (0.0587–0.0923) | 0.0791 (0.0596–0.1024) | 0.0715 (0.0005–0.1630) |
| $FEV_1$%predicted | 0.0809 (0.0611–0.1017) | 0.0809 (0.0545–0.1120) | 0.0380 (-0.0449–0.1233) |
| Spiro-metric LogReg | 0.0358 (0.0222–0.0514) | 0.0609 (0.0404–0.0843) | 0.0637 (-0.0053–0.1603) |
| Spiro-metric MLP | 0.0257 (0.0120–0.0394) | 0.0446 (0.0246–0.0704) | 0.0047 (-0.1067–0.1027) |
| Flow-volume MLP | 0.0160 (0.0042–0.0293) | 0.0107 (-0.0147–0.0375) | 0.0031 (-0.0782–0.0605) |

**Supplementary Table 4**: **Paired bootstrapping between the flow-volume ResNet18 and other candidate COPD models' AUC, AUPRC, and F1 scores across evaluation medical-record-based COPD disease status, future COPD-related hospitalization, and COPD-related death.** These results show the relative significance level of improvement of the flow-volume ResNet18 model over other candidate models from Supplementary Table 3 using paired bootstrapping ($n = 100$ bootstrapping trials).

| Model | Spearman R | Pearson R | Pearson R Squared |
|---|---|---|---|
| $FEV_1$/FVC Ratio | 0.1744 (0.1674–0.1815) | 0.2187 (0.2072–0.2310) | 0.0479 (0.0429–0.0533) |
| $FEV_1$%predicted | 0.1767 (0.1685–0.1843) | 0.1938 (0.1850–0.2047) | 0.0376 (0.0342–0.0419) |
| Spiro-metric LogReg | 0.1905 (0.1843–0.1983) | 0.2758 (0.2554–0.3026) | 0.0762 (0.0652–0.0915) |
| Spiro-metric MLP | 0.1964 (0.1907–0.2033) | 0.3158 (0.2960–0.3361) | 0.0998 (0.0876–0.1130) |
| Flow-volume MLP | 0.1988 (0.1928–0.2062) | 0.3366 (0.3177–0.3593) | 0.1134 (0.1009–0.1291) |
| Flow-volume ResNet18 | 0.2037 (0.1975–0.2114) | 0.3498 (0.3318–0.3703) | 0.1225 (0.1101–0.1371) |

**Supplementary Table 5**: **Comparison of model COPD risk with an individual's number of exacerbatory events.**

| Model | Spearman R | Pearson R | Pearson R Squared |
|---|---|---|---|
| $FEV_1/FVC$ Ratio | 0.0292 (0.0258–0.0335) | 0.1311 (0.1212–0.1421) | 0.0746 (0.0665–0.0853) |
| $FEV_1$%predicted | 0.0269 (0.0222–0.0323) | 0.1560 (0.1410–0.1745) | 0.0849 (0.0742–0.0985) |
| Spiro-metric LogReg | 0.0131 (0.0099–0.0156) | 0.0740 (0.0626–0.0848) | 0.0463 (0.0386–0.0533) |
| Spiro-metric MLP | 0.0072 (0.0049–0.0093) | 0.0340 (0.0254–0.0413) | 0.0227 (0.0169–0.0284) |
| Flow-volume MLP | 0.0049 (0.0028–0.0073) | 0.0132 (0.0078–0.0193) | 0.0091 (0.0053–0.0134) |

**Supplementary Table 6**: **Comparison of model COPD risk with an individual's number of exacerbatory events.** These results are similar to Supplementary Table 5 while to compute the significant level improvement between two models we used the paired bootstrapping ($n = 100$ bootstrapping trials).

See the attached Excel table.

**Supplementary Table 7**: **Comparison of cross-fold models.**

| Phenotype | Prevalence | #Hits | #Loci | SNP-heritability | Notes |
|---|---|---|---|---|---|
| ML-based COPD | NA | 796 | 356 | 0.2028 (0.0104) | |
| Proxy-GOLD | 7.22% | 45 | 30 | 0.0414 (0.0039) | |
| COPD (Medical-record-based) | 4.44% | 2 | 2 | 0.0112 (0.0029) | |
| COPD (Sakornsakolpat et al. 2019) | 13.86% | 117 | 84 | 0.0686 (0.0058) | |
| $FEV_1$ UKB (Shrine et al. 2019) | NA | 406 | 244 | 0.1799 (0.0073) | |
| FVC UKB (Shrine et al. 2019) | NA | 342 | 228 | 0.1779 (0.0063) | |
| $FEV_1/FVC$ UKB (Shrine et al. 2019) | NA | 669 | 328 | 0.2035 (0.01) | |
| Binarized ML-based COPD Sakornsakolpat et al. 2019 prevalence | 13.86% | 142 | 89 | 0.0726 (0.0054) | We fixed the prevalence of ML-based COPD to be the same as Sakornsakolpat et al. |
| Binarized ML-based COPD Proxy-GOLD prevalence | 7.22% | 56 | 37 | 0.0506 (0.0041) | We fixed the prevalence of ML-based COPD to be the same as Proxy-GOLD |

**Supplementary Table 8**: **Comparison of different model GWAS results and prevalence.**

See the attached Excel table.

**Supplementary Table 9**: **ML-based COPD GWS hits.** CHR, chromosome; POS, base-pair variant position; EA, effect allele; NEA, non-effect allele; BETA, estimated effect size; SE, standard error; SRC, imputed or genotyped variant; INFO, imputation INFO score (set to 1 for genotyped variants); P, GWAS p-value. GENE_CONTEXT, genomic context of the variant. Notation for gene context:

- Overlapping gene(s):
  - `[A]`: variant overlaps gene `A`
  - `[A,B]`: variant overlaps genes `A` and `B`
- Downstream genes:
  - `[]A`: variant position is $0 < p \leq 10^3$ bp upstream of closest downstream gene `A`
  - `[]-A`: variant position is $10^3 < p \leq 10^4$ bp upstream of closest downstream gene `A`
  - `[]-A`: variant position is $10^4 < p \leq 10^5$ bp upstream of closest downstream gene `A`
  - `[]--A`: variant position is $10^5 < p \leq 10^6$ bp upstream of closest downstream gene `A`
  - `[]`: closest downstream gene is further than $10^6$ bp
- Upstream genes: mirrors downstream gene notation, e.g., `B-[]` means variant position is $10^3 < p \leq 10^4$ bp downstream of closest gene `B`.

See the attached Excel table.

**Supplementary Table 10**: **ML-based COPD GWS loci.** CHR, chromosome; POS, base-pair variant position; EA, effect allele; NEA, non-effect allele; BETA, estimated effect size; SE, standard error; SRC, imputed or genotyped variant; INFO, imputation INFO score (set to 1 for genotyped variants); P, GWAS p-value. GENE_CONTEXT, genomic context of the variant. Notation for gene context:

- Overlapping gene(s):
  - `[A]`: variant overlaps gene `A`
  - `[A,B]`: variant overlaps genes `A` and `B`
- Downstream genes:
  - `[]A`: variant position is $0 < p \leq 10^3$ bp upstream of closest downstream gene `A`
  - `[]-A`: variant position is $10^3 < p \leq 10^4$ bp upstream of closest downstream gene `A`
  - `[]-A`: variant position is $10^4 < p \leq 10^5$ bp upstream of closest downstream gene `A`
  - `[]--A`: variant position is $10^5 < p \leq 10^6$ bp upstream of closest downstream gene `A`
  - `[]`: closest downstream gene is further than $10^6$ bp
- Upstream genes: mirrors downstream gene notation, e.g., `B-[]` means variant position is $10^3 < p \leq 10^4$ bp downstream of closest gene `B`.

See the attached Excel table.

**Supplementary Table 11**: **Novel ML-based COPD GWS hits.** CHR, chromosome; POS, base-pair variant position; EA, effect allele; NEA, non-effect allele; BETA, estimated effect size; SE, standard error; SRC, imputed or genotyped variant; INFO, imputation INFO score (set to 1 for genotyped variants); P, GWAS p-value. GENE_CONTEXT, genomic context of the variant. Notation for gene context:

- Overlapping gene(s):
  - `[A]`: variant overlaps gene `A`
  - `[A,B]`: variant overlaps genes `A` and `B`
- Downstream genes:
  - `[]A`: variant position is $0 < p \leq 10^3$ bp upstream of closest downstream gene `A`
  - `[]-A`: variant position is $10^3 < p \leq 10^4$ bp upstream of closest downstream gene `A`
  - `[]-A`: variant position is $10^4 < p \leq 10^5$ bp upstream of closest downstream gene `A`
  - `[]--A`: variant position is $10^5 < p \leq 10^6$ bp upstream of closest downstream gene `A`
  - `[]`: closest downstream gene is further than $10^6$ bp
- Upstream genes: mirrors downstream gene notation, e.g., `B-[]` means variant position is $10^3 < p \leq 10^4$ bp downstream of closest gene `B`.

See the attached Excel table.

**Supplementary Table 12**: **ML-based COPD GWS hits compared to other GWAS.** CHR, chromosome; POS, base-pair variant position; EA, effect allele; NEA, non-effect allele; BETA, estimated effect size; SE, standard error; SRC, imputed or genotyped variant; INFO, imputation INFO score (set to 1 for genotyped variants); P_ML-based COPD, ML-based COPD GWAS P-value; P_DeepNull, ML-based COPD GWAS P-value via DeepNull; P_FEV1/FVC, FEV1/FVC GWAS P-value; P_GOLD 2-4, GOLD 2-4 GWAS P-value; P_Sakornsakolpat 2019; COPD GWAS P-value obtained from Sakornsakolpat 2019 [S12]. GENE_CONTEXT, genomic context of the variant. Notation for gene context:

- Overlapping gene(s):
  - `[A]`: variant overlaps gene `A`
  - `[A,B]`: variant overlaps genes `A` and `B`
- Downstream genes:
  - `[]A`: variant position is $0 < p \leq 10^3$ bp upstream of closest downstream gene `A`
  - `[]-A`: variant position is $10^3 < p \leq 10^4$ bp upstream of closest downstream gene `A`
  - `[]-A`: variant position is $10^4 < p \leq 10^5$ bp upstream of closest downstream gene `A`
  - `[]--A`: variant position is $10^5 < p \leq 10^6$ bp upstream of closest downstream gene `A`
  - `[]`: closest downstream gene is further than $10^6$ bp
- Upstream genes: mirrors downstream gene notation, e.g., `B-[]` means variant position is $10^3 < p \leq 10^4$ bp downstream of closest gene `B`.

See the attached Excel table.

**Supplementary Table 13**: **Novel ML-based COPD GWS loci.** CHR, chromosome; POS, base-pair variant position; EA, effect allele; NEA, non-effect allele; BETA, estimated effect size; SE, standard error; SRC, imputed or genotyped variant; INFO, imputation INFO score (set to 1 for genotyped variants); P, GWAS p-value. GENE_CONTEXT, genomic context of the variant. Notation for gene context:

- Overlapping gene(s):
  - `[A]`: variant overlaps gene `A`
  - `[A,B]`: variant overlaps genes `A` and `B`
- Downstream genes:
  - `[]A`: variant position is $0 < p \leq 10^3$ bp upstream of closest downstream gene `A`
  - `[]-A`: variant position is $10^3 < p \leq 10^4$ bp upstream of closest downstream gene `A`
  - `[]-A`: variant position is $10^4 < p \leq 10^5$ bp upstream of closest downstream gene `A`
  - `[]--A`: variant position is $10^5 < p \leq 10^6$ bp upstream of closest downstream gene `A`
  - `[]`: closest downstream gene is further than $10^6$ bp
- Upstream genes: mirrors downstream gene notation, e.g., `B-[]` means variant position is $10^3 < p \leq 10^4$ bp downstream of closest gene `B`.

See the attached Excel table.

**Supplementary Table 14**: **ML-based COPD GWS Loci compared to other GWAS.** CHR, chromosome; POS, base-pair variant position; EA, effect allele; NEA, non-effect allele; BETA, estimated effect size; SE, standard error; SRC, imputed or genotyped variant; INFO, imputation INFO score (set to 1 for genotyped variants); P_ML-based COPD, ML-based COPD GWAS P-value; P_DeepNull, ML-based COPD GWAS P-value via DeepNull; P_FEV1/FVC, FEV1/FVC GWAS P-value; P_GOLD 2-4, GOLD 2-4 GWAS P-value; P_Sakornsakolpat 2019; COPD GWAS P-value obtained from Sakornsakolpat 2019 [S12].

See the attached Excel table.

**Supplementary Table 15**: **ML-based COPD GWS hits conditional on $FEV_1$/FVC.** CHR, chromosome; POS, base-pair variant position; EA, effect allele; NEA, non-effect allele; BETA, estimated effect size; SE, standard error; SRC, imputed or genotyped variant; INFO, imputation INFO score (set to 1 for genotyped variants); P, ML-based COPD GWAS P-value. GENE_CONTEXT, genomic context of the variant. Notation for gene context:

- Overlapping gene(s):
  - `[A]`: variant overlaps gene `A`
  - `[A,B]`: variant overlaps genes `A` and `B`
- Downstream genes:
  - `[]A`: variant position is $0 < p \leq 10^3$ bp upstream of closest downstream gene `A`
  - `[]-A`: variant position is $10^3 < p \leq 10^4$ bp upstream of closest downstream gene `A`
  - `[]-A`: variant position is $10^4 < p \leq 10^5$ bp upstream of closest downstream gene `A`
  - `[]--A`: variant position is $10^5 < p \leq 10^6$ bp upstream of closest downstream gene `A`
  - `[]`: closest downstream gene is further than $10^6$ bp
- Upstream genes: mirrors downstream gene notation, e.g., `B-[]` means variant position is $10^3 < p \leq 10^4$ bp downstream of closest gene `B`.

See the attached Excel table.

**Supplementary Table 16**: **ML-based COPD GWS loci conditional on $FEV_1$/FVC, $FEV_1$, FVC, and PEF.** CHR, chromosome; POS, base-pair variant position; EA, effect allele; NEA, non-effect allele; BETA, estimated effect size; SE, standard error; SRC, imputed or genotyped variant; INFO, imputation INFO score (set to 1 for genotyped variants); P, ML-based COPD GWAS P-value. GENE_CONTEXT, genomic context of the variant. Notation for gene context:

- Overlapping gene(s):
  - `[A]`: variant overlaps gene `A`
  - `[A,B]`: variant overlaps genes `A` and `B`
- Downstream genes:
  - `[]A`: variant position is $0 < p \leq 10^3$ bp upstream of closest downstream gene `A`
  - `[]-A`: variant position is $10^3 < p \leq 10^4$ bp upstream of closest downstream gene `A`
  - `[]-A`: variant position is $10^4 < p \leq 10^5$ bp upstream of closest downstream gene `A`
  - `[]--A`: variant position is $10^5 < p \leq 10^6$ bp upstream of closest downstream gene `A`
  - `[]`: closest downstream gene is further than $10^6$ bp
- Upstream genes: mirrors downstream gene notation, e.g., `B-[]` means variant position is $10^3 < p \leq 10^4$ bp downstream of closest gene `B`.

See the attached Excel table.

**Supplementary Table 17: ML-based COPD GWS hits conditional on $FEV_1/FVC$, FEV1, FVC, and PEF.** CHR, chromosome; POS, base-pair variant position; EA, effect allele; NEA, non-effect allele; BETA, estimated effect size; SE, standard error; SRC, imputed or genotyped variant; INFO, imputation INFO score (set to 1 for genotyped variants); P, ML-based COPD GWAS P-value. GENE_CONTEXT, genomic context of the variant. Notation for gene context:

- Overlapping gene(s):
  - `[A]`: variant overlaps gene `A`
  - `[A,B]`: variant overlaps genes `A` and `B`
- Downstream genes:
  - `[]A`: variant position is $0 < p \leq 10^3$ bp upstream of closest downstream gene `A`
  - `[]-A`: variant position is $10^3 < p \leq 10^4$ bp upstream of closest downstream gene `A`
  - `[]-A`: variant position is $10^4 < p \leq 10^5$ bp upstream of closest downstream gene `A`
  - `[]--A`: variant position is $10^5 < p \leq 10^6$ bp upstream of closest downstream gene `A`
  - `[]`: closest downstream gene is further than $10^6$ bp
- Upstream genes: mirrors downstream gene notation, e.g., `B-[]` means variant position is $10^3 < p \leq 10^4$ bp downstream of closest gene `B`.

See the attached Excel table.

**Supplementary Table 18: ML-based COPD GWS loci conditional on $FEV_1/FVC$**. CHR, chromosome; POS, base-pair variant position; EA, effect allele; NEA, non-effect allele; BETA, estimated effect size; SE, standard error; SRC, imputed or genotyped variant; INFO, imputation INFO score (set to 1 for genotyped variants); P, ML-based COPD GWAS P-value. GENE_CONTEXT, genomic context of the variant. Notation for gene context:

- Overlapping gene(s):
  - `[A]`: variant overlaps gene `A`
  - `[A,B]`: variant overlaps genes `A` and `B`
- Downstream genes:
  - `[]A`: variant position is $0 < p \leq 10^3$ bp upstream of closest downstream gene `A`
  - `[]-A`: variant position is $10^3 < p \leq 10^4$ bp upstream of closest downstream gene `A`
  - `[]-A`: variant position is $10^4 < p \leq 10^5$ bp upstream of closest downstream gene `A`
  - `[]--A`: variant position is $10^5 < p \leq 10^6$ bp upstream of closest downstream gene `A`
  - `[]`: closest downstream gene is further than $10^6$ bp
- Upstream genes: mirrors downstream gene notation, e.g., `B-[]` means variant position is $10^3 < p \leq 10^4$ bp downstream of closest gene `B`.

See the attached Excel table.

**Supplementary Table 19**: **ML-based COPD GWS hits via DeepNull.** CHR, chromosome; POS, base-pair variant position; EA, effect allele; NEA, non-effect allele; BETA, estimated effect size; SE, standard error; SRC, imputed or genotyped variant; INFO, imputation INFO score (set to 1 for genotyped variants); P, ML-based COPD GWAS P-value. GENE_CONTEXT, genomic context of the variant. Notation for gene context:

- Overlapping gene(s):
  - `[A]`: variant overlaps gene `A`
  - `[A,B]`: variant overlaps genes `A` and `B`
- Downstream genes:
  - `[]A`: variant position is $0 < p \leq 10^3$ bp upstream of closest downstream gene `A`
  - `[]-A`: variant position is $10^3 < p \leq 10^4$ bp upstream of closest downstream gene `A`
  - `[]-A`: variant position is $10^4 < p \leq 10^5$ bp upstream of closest downstream gene `A`
  - `[]--A`: variant position is $10^5 < p \leq 10^6$ bp upstream of closest downstream gene `A`
  - `[]`: closest downstream gene is further than $10^6$ bp
- Upstream genes: mirrors downstream gene notation, e.g., `B-[]` means variant position is $10^3 < p \leq 10^4$ bp downstream of closest gene `B`.

See the attached Excel table.

**Supplementary Table 20**: **ML-based COPD GWS loci via DeepNull.** CHR, chromosome; POS, base-pair variant position; EA, effect allele; NEA, non-effect allele; BETA, estimated effect size; SE, standard error; SRC, imputed or genotyped variant; INFO, imputation INFO score (set to 1 for genotyped variants); P, ML-based COPD GWAS P-value. GENE_CONTEXT, genomic context of the variant. Notation for gene context:

- Overlapping gene(s):
  - `[A]`: variant overlaps gene `A`
  - `[A,B]`: variant overlaps genes `A` and `B`
- Downstream genes:
  - `[]A`: variant position is $0 < p \leq 10^3$ bp upstream of closest downstream gene `A`
  - `[]-A`: variant position is $10^3 < p \leq 10^4$ bp upstream of closest downstream gene `A`
  - `[]-A`: variant position is $10^4 < p \leq 10^5$ bp upstream of closest downstream gene `A`
  - `[]--A`: variant position is $10^5 < p \leq 10^6$ bp upstream of closest downstream gene `A`
  - `[]`: closest downstream gene is further than $10^6$ bp
- Upstream genes: mirrors downstream gene notation, e.g., `B-[]` means variant position is $10^3 < p \leq 10^4$ bp downstream of closest gene `B`.

See the attached Excel table.

**Supplementary Table 21**: **ML-based COPD GWS DeepNull hits not significant in the $\mathbf{FEV_1/FVC}$ ratio GWAS.** CHR, chromosome; POS, base-pair variant position; EA, effect allele; NEA, non-effect allele; BETA, estimated effect size; SE, standard error; SRC, imputed or genotyped variant; INFO, imputation INFO score (set to 1 for genotyped variants); P, ML-based COPD GWAS P-value. GENE_CONTEXT, genomic context of the variant. Notation for gene context:

- Overlapping gene(s):
  - `[A]`: variant overlaps gene `A`
  - `[A,B]`: variant overlaps genes `A` and `B`
- Downstream genes:
  - `[]A`: variant position is $0 < p \leq 10^3$ bp upstream of closest downstream gene `A`
  - `[]-A`: variant position is $10^3 < p \leq 10^4$ bp upstream of closest downstream gene `A`
  - `[]-A`: variant position is $10^4 < p \leq 10^5$ bp upstream of closest downstream gene `A`
  - `[]--A`: variant position is $10^5 < p \leq 10^6$ bp upstream of closest downstream gene `A`
  - `[]`: closest downstream gene is further than $10^6$ bp
- Upstream genes: mirrors downstream gene notation, e.g., `B-[]` means variant position is $10^3 < p \leq 10^4$ bp downstream of closest gene `B`.

See the attached Excel table.

**Supplementary Table 22**: **ML-based COPD GWS DeepNull loci not significant in the $\mathbf{FEV_1/FVC}$ ratio GWAS.** CHR, chromosome; POS, base-pair variant position; EA, effect allele; NEA, non-effect allele; BETA, estimated effect size; SE, standard error; SRC, imputed or genotyped variant; INFO, imputation INFO score (set to 1 for genotyped variants); P, ML-based COPD GWAS P-value. GENE_CONTEXT, genomic context of the variant. Notation for gene context:

- Overlapping gene(s):
  - `[A]`: variant overlaps gene `A`
  - `[A,B]`: variant overlaps genes `A` and `B`
- Downstream genes:
  - `[]A`: variant position is $0 < p \leq 10^3$ bp upstream of closest downstream gene `A`
  - `[]-A`: variant position is $10^3 < p \leq 10^4$ bp upstream of closest downstream gene `A`
  - `[]-A`: variant position is $10^4 < p \leq 10^5$ bp upstream of closest downstream gene `A`
  - `[]--A`: variant position is $10^5 < p \leq 10^6$ bp upstream of closest downstream gene `A`
  - `[]`: closest downstream gene is further than $10^6$ bp
- Upstream genes: mirrors downstream gene notation, e.g., `B-[]` means variant position is $10^3 < p \leq 10^4$ bp downstream of closest gene `B`.

| Phenotype 1 | Phenotype 2 | $r_g$ |
|---|---|---|
| ML-based COPD | COPD (Sakornsakolpat et al. 2019) | 0.8985 (0.0736) |
| ML-based COPD | $FEV_1$ UKB (Shrine et al. 2019) | -0.7031 (0.0441) |
| ML-based COPD | $FEV_1$/FVC UKB (Shrine et al. 2019) | -0.7837 (0.0491) |
| ML-based COPD | FVC UKB (Shrine et al. 2019) | -0.3440 (0.0261) |
| ML-based COPD | PEF UKB (Shrine et al. 2019) | -0.5757 (0.0385) |
| ML-based COPD | Height UKB | 0.0580 (0.0214) |
| ML-based COPD | Asthma UKB | 0.3980 (0.037) |
| COPD (Sakornsakolpat et al. 2019) | Asthma UKB | 0.4815 (0.0531) |
| COPD (Sakornsakolpat et al. 2019) | Height UKB | 0.1289 (0.0329) |

**Supplementary Table 23**: **COPD genetic correlation with existing GWAS.** The genetic correlation between two phenotypes is computed using S-LDSC.

See the attached Excel table.

**Supplementary Table 24**: **ML-based COPD novel loci replication in GBMI, ICGC, and SpiroMeta.** We analyzed three studies that do not include UK Biobank samples to quantify loci replication. These three datasets are GBMI (Global Biobank Meta-analysis Initiative) [S32], SpiroMeta [S33], and ICGC (International COPD Genetics Consortium) [S11]. CHR, chromosome; POS, base-pair variant position; EA, effect allele; NEA, non-effect allele; BETA, estimated effect size; SE, standard error; ICGC-EA, ICGC-BETA, ICGC-P, GMBI-P, SpiroMeta-FVC-EA, SpiroMeta-FVC-BETA, SpiroMeta-FVC-P, SpiroMeta-FEV1-EA, SpiroMeta-FEV1-BETA, SpiroMeta-FEV1-P, SpiroMeta-Ratio-EA, SpiroMeta-Ratio-BETA, SpiroMeta-Ratio-P, Strict Replication, and Supportive Replication. we defined *supportive* replication as consistent effect size direction across studies. The ICGC and GBMI GWAS are based on a COPD phenotype; thus, we expect their effect size signs to match our ML-based COPD. SpiroMeta phenotypes, on the other hand, capture lung function, so we expect their effect size signs to be the opposite of our ML-based COPD signs. We defined *strict* replication as consistent effect size direction in any study with Bonferroni correction of $P < 0.1$ (one-sided) for that study.

See the attached Excel table.

**Supplementary Table 25**: **Cell-type specific chromatin marks identify disease-relevant cell-type for ML-based COPD**. We performed cell-type specific chromatin heritability enrichment analysis via S-LDSC. We obtained the cell-type specific chromatin from S-LDSC website on BROAD. The second column is the regression coefficient corresponding to the cell type specific annotation and third column is the corresponding standard mean error. The last column is the p-value from a one-sided test that the regression coefficient is greater than zero. It is recommended by the authors [S60] that significant level of regression coefficient (e.g., p-value column) should be used to identify disease-relevant cell-type.

See the attached Excel table.

**Supplementary Table 26**: **S-LDSC enrichments of ML-based COPD for cell-type specific chromatin marks.** In addition to detecting the COPD-relevant cell-type, we compute the enrichment of ML-based COPD in cell-type specific chromatin marks. `Prop._SNPs` is the fraction of SNPs that is covered by each cell-type specific annotation. `Prop._h2` is the fraction of ML-based COPD that is captured by each cell-type specific annotation. `Enrichment` is defined as the $\frac{\texttt{Prop.\_h2}}{\texttt{Prop.\_SNPs}}$. `Enrichment_std_error` is the standard error of enrichment computed by S-LDSC via block-jackknife.

See the attached Excel table.

**Supplementary Table 27**: **Cell-type specific chromatin marks identify disease-relevant cell-type for ML-based COPD conditional on $FEV_1$/FVC** . We performed cell-type specific chromatin heritability enrichment analysis via S-LDSC.

| Term | Description | Region P | Gene P | Num regions |
|---|---|---|---|---|
| GO:0072359 | circulatory system development | 4.49e-05 | 2.62e-12 | 78 |
| GO:0007507 | heart development | 1.37e-04 | 1.61e-10 | 56 |
| MP:0005385 | cardiovascular system phenotype | 1.49e-04 | 1.35e-09 | 137 |
| HP:0001626 | Abnormality of the cardiovascular system | 7.61e-04 | 7.53e-04 | 71 |
| MP:0002127 | abnormal cardiovascular system morphology | 8.33e-04 | 2.82e-08 | 111 |
| MP:0001544 | abnormal cardiovascular system physiology | 1.29e-03 | 6.25e-09 | 97 |
| MP:0001175 | abnormal lung morphology | 5.76e-03 | 1.67e-05 | 47 |
| GO:0003007 | heart morphogenesis | 5.80e-03 | 4.21e-07 | 34 |
| MP:0001176 | abnormal lung development | 2.34e-02 | 1.33e-03 | 21 |
| MP:0003115 | abnormal respiratory system development | 2.44e-02 | 3.32e-04 | 23 |
| HP:0030680 | Abnormality of cardiovascular system morphology | 3.33e-02 | 4.65e-03 | 52 |
| MP:0002132 | abnormal respiratory system morphology | 3.90e-02 | 1.77e-05 | 60 |
| GO:0003205 | cardiac chamber development | 4.71e-02 | 1.09e-06 | 27 |
| GO:0003279 | cardiac septum development | 4.78e-02 | 8.32e-07 | 22 |

**Supplementary Table 28**: **Cardiovascular and respiratory term enrichments of the ML-based COPD loci.** Enrichments were computed using GREAT [S35] with default parameters. The 82 total terms significant at Bonferroni-corrected P-value 0.05 by both the region-based binomial and gene-based hypergeometric tests were filtered to those with description that matched the regular expression 'cardiac|cardio|cardial|heart|circulatory|respir|lung|pulmon'.

| Ground Truth | PRS | AUROC | AUPRC | Top Decile Prevalence | R |
|---|---|---|---|---|---|
| Eval. MRB COPD | ResNet18 | 0.550 (0.541–0.560) | 0.088 (0.084–0.092) | 0.097 (0.089–0.104) | 0.046 (0.037–0.056) |
| Eval. MRB COPD | MRB COPD | 0.517 (0.509–0.528) | 0.079 (0.076–0.082) | 0.081 (0.076–0.086) | 0.018 (0.010–0.027) |
| Eval. MRB COPD | Sakornsakolpat | 0.538 (0.529–0.548) | 0.085 (0.081–0.089) | 0.095 (0.086–0.103) | 0.036 (0.028–0.045) |
| Eval. MRB COPD | $FEV_1$/FVC | 0.540 (0.531–0.551) | 0.085 (0.082–0.090) | 0.095 (0.088–0.104) | 0.037 (0.028–0.047) |
| Hospitalization | ResNet18 | 0.564 (0.549–0.577) | 0.023 (0.021–0.025) | 0.025 (0.022–0.028) | 0.029 (0.022–0.036) |
| Hospitalization | MRB COPD | 0.514 (0.504–0.526) | 0.018 (0.017–0.020) | 0.020 (0.018–0.021) | 0.006 (0.001–0.013) |
| Hospitalization | Sakornsakolpat | 0.551 (0.537–0.565) | 0.022 (0.020–0.024) | 0.025 (0.022–0.028) | 0.024 (0.018–0.031) |
| Hospitalization | $FEV_1$/FVC | 0.560 (0.546–0.573) | 0.023 (0.021–0.025) | 0.026 (0.023–0.029) | 0.028 (0.021–0.034) |
| Death | ResNet18 | 0.598 (0.557–0.632) | 0.004 (0.003–0.005) | 0.004 (0.003–0.005) | 0.016 (0.009–0.023) |
| Death | MRB COPD | 0.503 (0.473–0.537) | 0.002 (0.002–0.003) | 0.002 (0.002–0.003) | 0.001 (-0.004–0.007) |
| Death | Sakornsakolpat | 0.575 (0.533–0.606) | 0.003 (0.002–0.004) | 0.003 (0.002–0.005) | 0.013 (0.006–0.019) |
| Death | $FEV_1$/FVC | 0.599 (0.554–0.645) | 0.004 (0.003–0.005) | 0.004 (0.003–0.005) | 0.016 (0.009–0.024) |

**Supplementary Table 29**: **Comparison of PRSs in UKB.** The PRSs are defined based on the GWAS effect sizes of ML-based COPD, Medical-record-based (MRB) COPD, and Sakornsakolpat et al. [S12], and the $FEV_1$/FVC. The numbers in the parenthesis show the 95% confidence interval. R is the Pearson correlation between true phenotypic values and estimated PRS.

| Ground Truth | PRS | AUROC | AUPRC | Top Decile Prevalence | R |
|---|---|---|---|---|---|
| COPD Status | ResNet18 | 0.615 (0.598–0.631) | 0.632 (0.615–0.650) | 0.682 (0.647–0.720) | 0.205 (0.178–0.232) |
| COPD Status | MRB COPD | 0.525 (0.511–0.538) | 0.550 (0.536–0.565) | 0.547 (0.517–0.577) | 0.046 (0.022–0.066) |
| COPD Status | Sakornsakolpat | 0.616 (0.599–0.630) | 0.623 (0.602–0.640) | 0.689 (0.655–0.721) | 0.202 (0.173–0.227) |
| COPD Status | $FEV_1/FVC$ | 0.618 (0.602–0.632) | 0.627 (0.608–0.644) | 0.688 (0.644–0.723) | 0.206 (0.182–0.230) |
| pctEmph_vida | ResNet18 | NA | NA | NA | 0.106 (0.079–0.127) |
| pctEmph_vida | MRB COPD | NA | NA | NA | 0.043 (0.017–0.066) |
| pctEmph_vida | Sakornsakolpat | NA | NA | NA | 0.140 (0.114–0.168) |
| pctEmph_vida | $FEV_1/FVC$ | NA | NA | NA | 0.138 (0.113–0.157) |
| Pi10_SRWA_vida | ResNet18 | NA | NA | NA | 0.051 (0.030–0.072) |
| Pi10_SRWA_vida | MRB COPD | NA | NA | NA | 0.000 (-0.023–0.024) |
| Pi10_SRWA_vida | Sakornsakolpat | NA | NA | NA | 0.021 (-0.005–0.042) |
| Pi10_SRWA_vida | $FEV_1/FVC$ | NA | NA | NA | 0.026 (-0.001–0.051) |

**Supplementary Table 30**: **Comparison of PRSs in COPDGene.** The PRSs are defined based on the GWAS effect sizes of ML-based COPD, Medical-record-based (MRB) COPD, and Sakornsakolpat et al. [S12], and the ratio, $FEV_1/FVC$. The numbers in the parenthesis show the 95% confidence interval. The effected individuals are defined as the individuals with final GOLD stage 2, 3, and 4 post-QA. R is the Pearson correlation between true phenotypic values and estimated PRS.

See the attached Excel table.

**Supplementary Table 31**: **PheWAS of ML-based COPD significant hits.** We extracted the statistical test for all phenotypes in Neal lab and FinnGen of all ML-based COPD significant hits (796 independent GWS hits). In the case of FinnGen, variants position used the GRCh38-hg38 build, we used USCS liftover to match the variants. VIP is the variant ID created by `chr:bp:ref:alt` where `chr` is the chromosome, `bp` is the variant position, `ref` is the reference allele, and `alt` is the alternative allele. P is the p-value obtrained from Neal lab and FinnGen GWAS and FDR is the computed false discovery rate.

See the attached Excel table.

**Supplementary Table 32**: **Summary of ML-based COPD PheWAS based on phenotypes**. We sorted phenotypes in Neal lab and FinnGen based on number significant hits obtained from ML-based COPD. We used the FDR $< 5\%$ to consider significant.

| Model | Medical-record-based AUC | Hospitalization AUC | Death AUC |
|---|---|---|---|
| $FEV_1$/FVC Ratio | 0.6977 (0.6622–0.7286) | 0.8225 (0.7542–0.8705) | 0.9862 (0.9732–0.9953) |
| Flow-volume ResNet18 | **0.7523 (0.7201–0.7890)** | **0.8705 (0.8262–0.9151)** | **0.9918 (0.9781–0.9992)** |

| Model | Medical-record-based AUPRC | Hospitalization AUPRC | Death AUPRC |
|---|---|---|---|
| $FEV_1$/FVC Ratio | 0.0914 (0.0708–0.1145) | 0.0383 (0.0230–0.0587) | 0.0357 (0.0107–0.0888) |
| Flow-volume ResNet18 | **0.1786 (0.1398–0.2190)** | **0.1031 (0.0616–0.1558)** | **0.2851 (0.0472–0.5777)** |

| Model | Medical-record-based F1 | Hospitalization F1 | Death F1 |
|---|---|---|---|
| $FEV_1$/FVC Ratio | 0.1866 (0.1491–0.2178) | 0.0898 (0.0588–0.1228) | 0.0507 (0.0077–0.1048) |
| Flow-volume ResNet18 | **0.2400 (0.1969–0.2856)** | **0.1915 (0.1259–0.2532)** | 0.1495 (0.0000–0.4369) |

**Supplementary Table 33**: **Comparison of model AUC, AUPRC, and F1 scores across tasks in non-Europeans.** Performance metrics are calculated using non-European individuals with valid spirometry blows for medical-record-based COPD disease status ($n = 8,201$, prevalence = 3.244%), future COPD-related hospitalization($n = 21,878$, prevalence = 0.384%), and COPD-related death($n = 26,125$, prevalence= 0.034%). $FEV_1$/FVC Ratio denotes a risk model based on standard spirometry metrics. The flow-volume ResNet18 model utilized the entire flow-volume curve. We performed hyperparameter sweeps for each class of deep learning model, selecting the model that minimized the binary cross entropy loss over the modeling validation set. 95% confidential intervals were generated using bootstrapping ($n = 100$ bootstrapping trials). Bold values denote that a model is statistically better than other models for the given task metric ($n = 100$ bootstrapping trials).

| Codes | Description |
|---|---|
| J41 | Simple and mucopurulent chronic bronchitis |
| J42 | Unspecified chronic bronchitis |
| J43 | Emphysema |
| J44 | Other chronic obstructive pulmonary disease |

**Supplementary Table 34**: **COPD ICD10 codes.**

| Dataset | Split | $n$ | MRB | Eval. MRB | GOLD | Hospitalization | Death |
|---|---|---|---|---|---|---|---|
| Modeling | Train | 259746 | 0.0383 | 0.0473 | 0.0725 | 0.0075 | 0.0007 |
| Modeling | Validation | 65281 | 0.0388 | 0.0479 | 0.0720 | 0.0072 | 0.0007 |
| Fold 1 | Train | 128739 | 0.0384 | 0.0486 | 0.0711 | 0.0076 | 0.0007 |
| Fold 1 | Validation | 32310 | 0.0391 | 0.0477 | 0.0719 | 0.0073 | 0.0008 |
| Fold 2 | Train | 129691 | 0.0379 | 0.0456 | 0.0734 | 0.0072 | 0.0007 |
| Fold 2 | Validation | 32637 | 0.0381 | 0.0480 | 0.0714 | 0.0069 | 0.0006 |
| PRS | Holdout | 110739 | 0.0640 | 0.0748 | - | 0.0053 | 0.0021 |

**Supplementary Table 35**: **Prevalence of COPD cases across datasets.** MRB stands for medical-record-based. GOLD labels are omitted for the PRS holdout set since it contains individuals with invalid blows. $n$ is the number of individuals in each dataset.

| Dataset | Split | $n$ | $FEV_1$ | FVC | $FEV_1/FVC$ | $FEV_1$%predicted |
|---|---|---|---|---|---|---|
| Modeling | Train | 259746 | $2.84 \pm 0.75$ | $3.76 \pm 0.95$ | $0.75 \pm 0.07$ | $96.47 \pm 16.96$ |
| Modeling | Validation | 65281 | $2.84 \pm 0.75$ | $3.76 \pm 0.94$ | $0.75 \pm 0.07$ | $96.48 \pm 16.90$ |
| Fold 1 | Train | 128739 | $2.84 \pm 0.75$ | $3.76 \pm 0.95$ | $0.75 \pm 0.07$ | $96.50 \pm 16.84$ |
| Fold 1 | Validation | 32310 | $2.84 \pm 0.75$ | $3.76 \pm 0.95$ | $0.75 \pm 0.07$ | $96.53 \pm 16.90$ |
| Fold 2 | Train | 129691 | $2.84 \pm 0.75$ | $3.76 \pm 0.95$ | $0.75 \pm 0.07$ | $96.47 \pm 17.04$ |
| Fold 2 | Validation | 32637 | $2.84 \pm 0.75$ | $3.76 \pm 0.94$ | $0.75 \pm 0.07$ | $96.47 \pm 16.86$ |

**Supplementary Table 36**: **Mean spirometry metrics across datasets.** The PRS holdout set is omitted since it contains only individuals with invalid blows. $n$ is the number of individuals in each dataset and $FEV_1$%predicted is the predicted $FEV_1$ using age, sex, and height as features.

See the attached Excel table.

**Supplementary Table 37**: **Overview of hyperparameters considered for model architectures.**

See the attached Excel table.

**Supplementary Table 38**: **Overview of the final hyperparameters used for each architecture.**

| Pheno Name | UKB Data-Field |
|---|---|
| Age | 21003 |
| BMI | 21001 |
| Sex | 31 |
| Height | 50 |
| Current tobacco smoking | 1239 |
| Past tobacco smoking | 1249 |
| Smoking pack years | 20161 |

**Supplementary Table 39**: **Set of non-PC covariates utilized in ML-based GWAS.** UKB Data-Field code for non-PC covariates.
